# Real-time diagnostics and personalized wound therapy powered by AI and bioelectronics

**DOI:** 10.1101/2025.01.12.25320338

**Authors:** Houpu Li, Hsin-ya Yang, Fan Lu, Wan Shen Hee, Narges Asefifeyzabadi, Prabhat Baniya, Anthony Gallegos, Kaelan Schorger, Kan Zhu, Cynthia Recendez, Maryam Tebyani, Manasa Kesapragada, Gordon Keller, Sujung Kim, George Luka, Ksenia Zlobina, Tiffany Nguyen, Sydnie Figuerres, Celeste Franco, Koushik Devarajan, Alexie Barbee, Kylie Lin, Shannon M. Clayton, Annabelle Eaton, Elham Aslankoohi, Athena M. Soulika, Min Zhao, Mircea Teodorescu, Marcella Gomez, Roslyn Rivkah Isseroff, Marco Rolandi

**Author notes:** These authors contributed equally to this work (Houpu Li, Hsin-ya Yang, Fan Lu, Wan Shen Hee).

## Abstract

Impaired wound healing affects millions worldwide, especially those without timely healthcare access. TheraHeal provides a portable, wireless platform for real-time, continuous, and personalized wound care. The platform integrates a wearable device for wound imaging and delivery of therapy with an AI Physician. The AI Physician analyzes wound images, diagnoses the wound stage, and prescribes therapies to guide optimal healing. Bioelectronic actuators in the wearable device deliver therapies, including electric fields or drugs, dynamically in a closed-loop system. TheraHeal evaluates wound progress, adjusts therapy as needed, and sends updates to human physicians through a graphical user interface, which also supports manual intervention. In a large animal model, TheraHeal enhanced tissue regeneration, reduced inflammation, and accelerated healing, showcasing its potential to transform personalized wound care.

## Introduction

The average person suffers 1–3 wounds annually, resulting in an estimated 24 billion wounds worldwide each year^1^. These wounds range from minor scrapes and traumatic injuries to surgical incisions and chronic ulcers^2^. While many wounds heal with basic at-home care, others require timely medical intervention to ensure proper healing^3^. Delays in access to healthcare can lead to complications, including scarring, permanent tissue damage, infection, sepsis, and even death ^4^. Wound type and individual variability further complicate the selection of optimal treatments^5^.

Traditional methods often rely on static, standardized protocols and assessments that cannot adapt to the individual patient, the type, and importantly, the changing state of a wound ^6^. These deficits can result in prolonged recovery, increased risks of complications, and less-than-ideal healing outcomes^7^.

Recent advances in wearables and smart bandages enable personalized wound treatment^8^ using remotely controlled microneedle^9, 10^, photo patch^11^, and wireless bandage with sensing and drug delivery function^12, 13^. This treatment can be optimized through continuous wound monitoring with onboard sensors to improve delivery and outcomes^12, 13^. However, wound healing is inherently complex, progressing through hemostasis, inflammation, proliferation, and maturation stages, each involving distinct cellular and biochemical processes that require therapy with specific biochemical signaling for optimal healing^14^. Incorporating sensors to monitor every biochemical process in the wound remains a challenge^15^. To address this, machine learning approaches have significantly improved healthcare by processing and inferring diagnostics from large, continuously updating datasets^16, 17^. Holistic, machine learning-driven methods can assess the overall wound stage, indicate required treatments, and adjust them in real time based on the wound trajectory^18, 19^. Here, we introduce TheraHeal a wireless, integrated platform for real-time, onboard, personalized wound theranostics, utilizing AI and machine learning to continuously determine the updated wound stages, prescribe and deliver therapies for optimized healing (Fig. 1).

**Fig.1.**
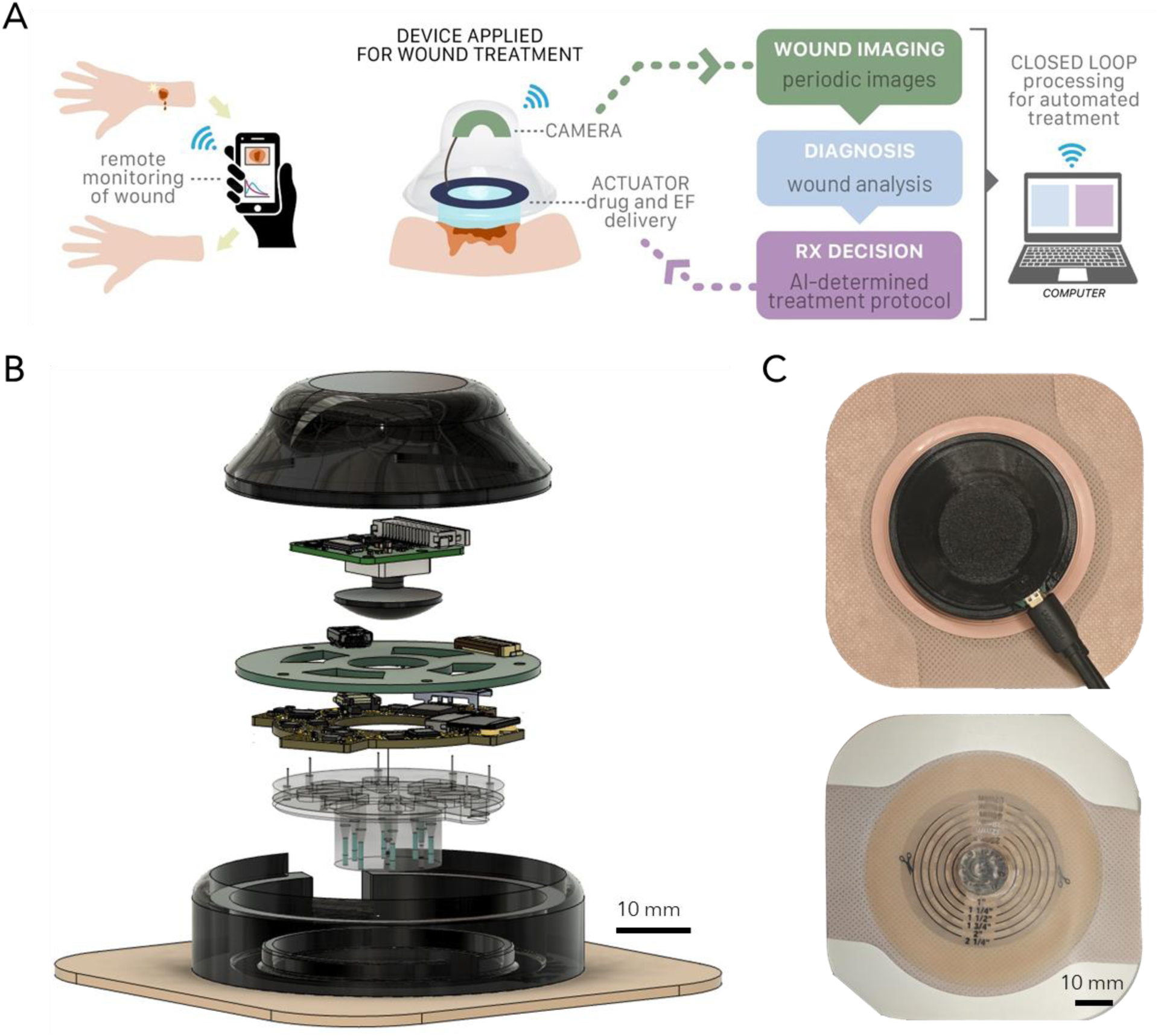
Wireless, integrated system for real-time, onboard, personalized wound theranostics. (A) There Heal platform. (B) CAD model of There Heal wearable device, illustrating the integrated camera for wound monitoring and actuator for drug delivery or stimulation. (C) Photograph of the TheraHeal wearable, demonstrating the final setup with the camera and actuator, highlighting the translation of the CAD model into a functional prototype and integration with bandage for ease of application.

## Results and discussion

### TheraHeal

The TheraHeal platform includes two main components: the TheraHeal wearable device (Fig. 1 and S1), which monitors wounds and delivers on-demand therapy, and the TheraHeal AI Physician, a machine learning (ML) driven intelligent diagnostic and decision-making algorithm with a graphical user interface (GUI) for human physician oversight, if needed (Figure 1A). The closed-loop theranostics process begins when the onboard camera of the TheraHeal wearable device captures an image of the wound and transmits it wirelessly to the TheraHeal AI Physician. The AI Physician analyzes the image, generates a diagnosis of wound stage, and prescribes a treatment plan to accelerate wound healing. The wearable device receives the prescribed therapy wirelessly and implements it using bioelectronic actuators that deliver an electric field or a drug of choice (Fig. 1A). After the therapy takes effect for a specified duration, the wearable device captures a new wound image and restarts the diagnostic and therapeutic cycle (Fig. 1A). The wearable TheraHeal includes a camera module, two printed circuit boards (PCBs) for camera illumination, onboard computing and wireless transmission, and a transparent polydimethylsiloxane (PDMS) body that houses reservoirs for therapy storage and the bioelectronic actuators (Fig. 1B). A 3D-printed waterproof enclosure encases these components and integrates a USB-C port for connection to an external power supply (Fig. 1C). This enclosure attaches directly to a commercially available bandage, originally developed for Colostomy, ensuring a convenient and secure way to keep the device in place during treatment (Fig. 1C). For this work, we test TheraHeal with a combination therapy of Electric Field (EF) and Fluoxetine (Prozac^TM^). EF is a well-known stimulant for healing wounds ^20^ ^21, 22^ by enhancing cell migration. We have previously demonstrated that both topical ^23^ and bioelectronic delivery of Fluoxetine (Flx) promotes healing in a small animal model^24^.

### Camera Module

Physicians rely on wound imaging for initial assessment as a critical tool for judging healing and guiding treatment decisions^25, 26^. The TheraHeal AI Physician mimics the diagnostic approach used by physicians, enabling the creation of personalized treatment regimens^27,28^. To achieve this functionality, the TheraHeal wearable device integrates an imaging module with a camera, a plano-convex lens, an illumination PCB, and a microcontroller (Fig. 2A and Fig. S2). The illumination PCB, designed as a circular ring with 12 LED banks, surrounds the camera to ensure consistent lighting (Fig. 2B). The optical path from the wound to the camera passes through a transparent PDMS layer (n=1.4), which incorporates the bioelectronic actuators and drug reservoirs^29^. A custom molding process ensures the PDMS achieves the optical clarity required for capturing high-quality wound images (Fig. 2C and Fig. S3 and S4). The imaging module captures 11 images in a z-stack to optimize image quality by focusing on different planes and accommodating non-uniform biological surfaces. After capturing the images, the module transmits them wirelessly to a designated receiver and initiates a 2-hour sleep mode before starting the next imaging cycle (Fig. 2D). The sleep mode is designed to minimize power usage and ensure that the batteries last at least 24hrs, the typical wound monitoring cycle. The controller architecture integrates a microcontroller powered by a 5V power bank, a CSI-2 multiplexer board, and shield boards (Fig. 2E). The microcontroller uses CSI-2 and GPIO/I2C interfaces to communicate with shield boards and links seamlessly with breakout boards, adapter boards, illumination PCBs, and imaging sensors through CSI-2 and HDMI connections (Fig. 2E). The module captures ex vivo images of swine skin (Fig. 2F) and in vivo subcutaneous wound images. These images guide machine-learning-based treatment algorithms and undergo post-processing to provide deeper insights into wound healing progression.

**Fig 2.**
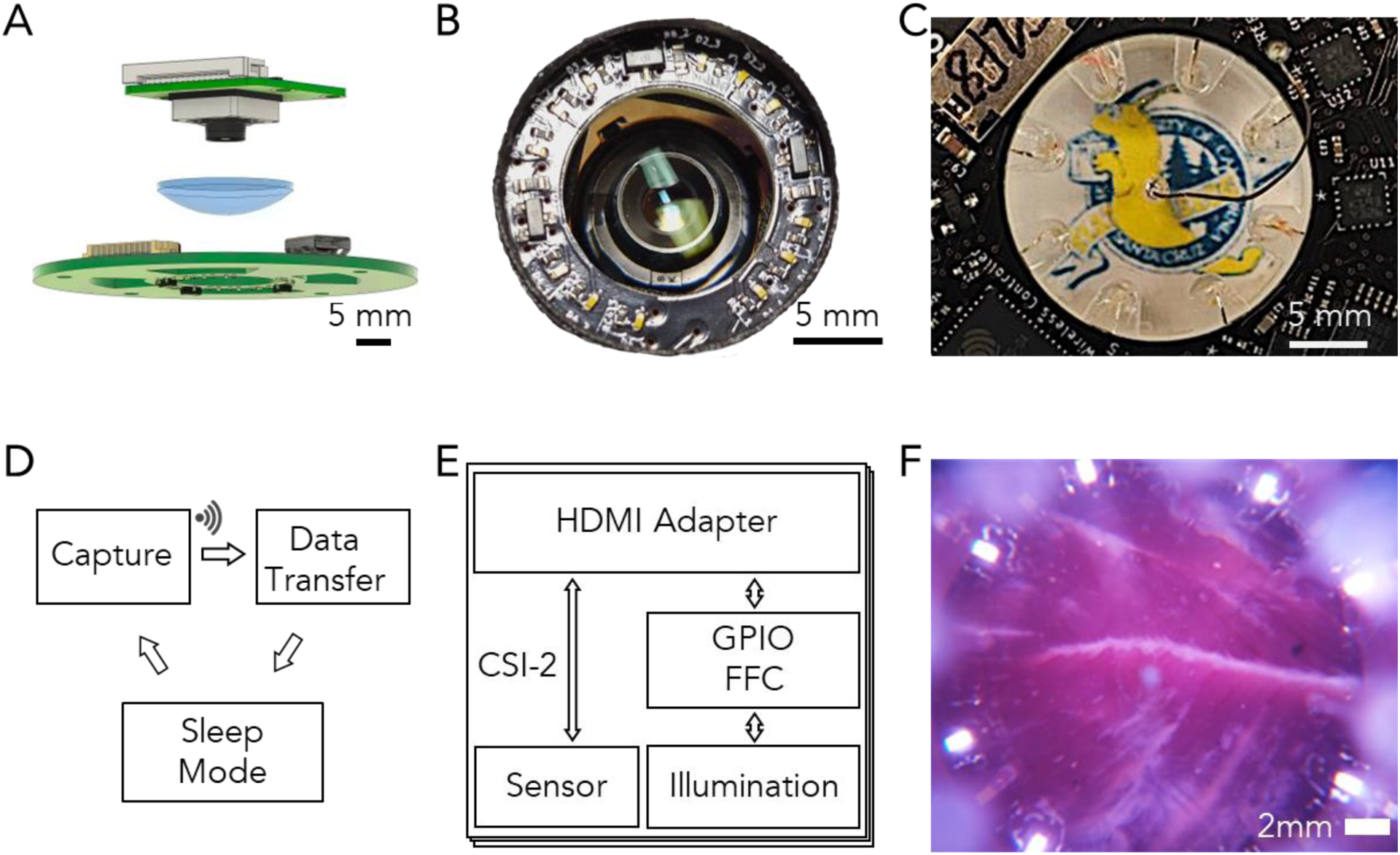
Overview of Imaging Subsystem and Key Components. Fig 2 Overview of Imaging Subsystem. (A) CAD of imaging components, (B) camera module with illumination board, (C) UCSC logo via PDMS optical path, (D) multi-day capture protocol, (E) controller communication via HDMI, CSI-2, GPIO, and I2C, (F) ex vivo porcine skin image from the device camera. optical path and the ion pump module device, and calibration target. (F) Sample wound image of an in vivo wound on a swine full thickness wound model.

### Bioelectronic Therapy Delivery

The ability to deliver the therapy on demand and optimize healing progression comes from the second part of the TheraHeal wearable, a set of bioelectronic actuators (Fig. 3). This component consists of two core parts: a ring-shaped PCB and an ion pump body, which features eight reservoirs and channels fabricated from PDMS (Fig. 3A). The PCB incorporates Wi-Fi connectivity, enabling the microcontroller to receive commands based on the AI Physician, which specifies voltages or currents for each channel (Fig. 3B). These commands are processed by the microcontroller, generating low-level electrical signals^30^. The onboard potentiostat routes the signals to specific channels, delivers them, and conducts electrochemical measurements, recording current as a function of time and applied voltage, V_EF_ for electric field and V_Flx_ for fluoxetine respectively(Fig. 3C). Recording current is important to estimate the electric field strength^22, 31^ and the delivered dose of fluoxetine ^24, 32, 33^. The ion pump body comprises reservoirs solution housing the individually addressable working electrodes and nine capillaries containing an anionic polymer designed to act as a filter for positive ions ^34^. These capillaries link the reservoirs to the wound bed (Fig. 3D). Capillaries were fabricated using a previously established method (Fig. S5-6)^34^. For EF delivery, the reservoirs are filled with physiologically compatible, sterile saline solution and equipped with Ag/AgCl electrodes functioning as the working and reference electrodes. The reference electrode is strategically positioned at the wound center, ensuring that the EF direction radiates inward from the wound edge to its center^21,22^. For fluoxetine delivery, reservoirs are filled with a solution of fluoxetine hydrochloride (Flx in HCl) at a concentration ensuring protonation and a net positive charge^35^. V_Flx_, drives Flx molecules from the reservoir’s working electrode into the wound bed (Fig. S9-10)^24^. For combined treatment, four reservoirs are dedicated to Ef delivery (blue), while the remaining four are used for Flx delivery (red) (Fig. 3D). With V_EF_= 4.7 V, the electric field reaches peaks of 400 mV/cm, oriented radially toward the wound center, as validated by COMSOL simulations (Fig. 3E, Fig S7, S8). With V_Flx_, the system delivers a controlled rate of Flx, with a concentration gradient—higher near the capillaries and lower at the wound center. The capillaries are positioned at the wound edge to create a clear optical path for the camera module to acquire wound images. The amount of Flx delivered is determined by measuring the current at each electrode (I_Flx_) and calibrating delivery efficiency using high-performance liquid chromatography (HPLC) ^36^(Fig. S9). Before starting the in vivo experiments, each device undergoes in vitro and ex vivo testing to confirm full functionality (Fig. S11-14).

**Fig. 3.**
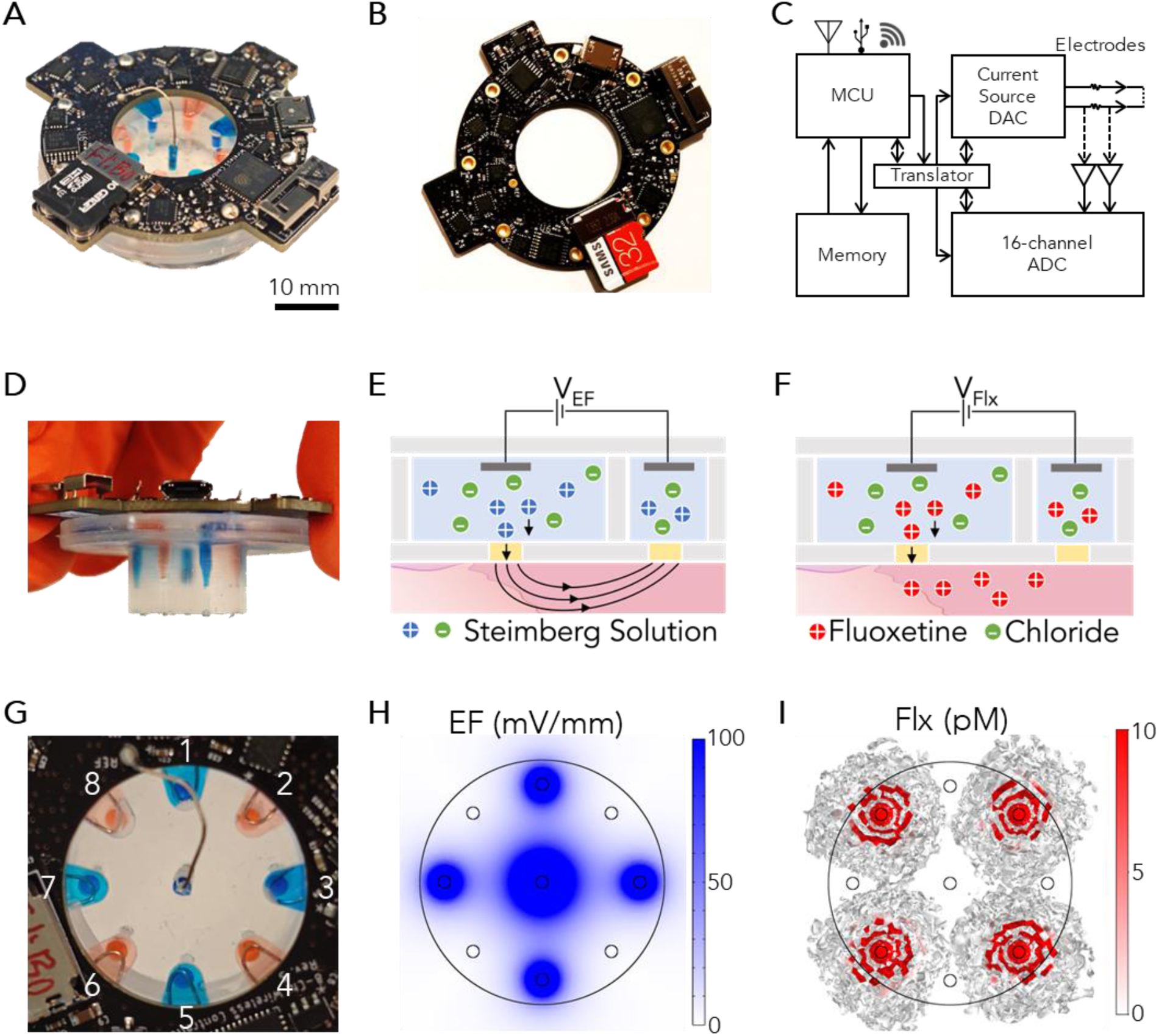
Setup of the Bioelectronic Actuator for electric field and iontophoretic drug delivery. (A) Actuator device photograph. (B) PCB with wireless potentiostat. (C) Microcontroller architecture, (D) Fluidic channel with dye solutions. (E) Fluoxetine delivery schematic. (F) Electric field delivery schematic. (G) Top view. (H) Fluoxetine dosage graph. (I) Electric field strength graph.

### AI Physician

The goal of the AI Physician is to shift a wound from a slow or suboptimal healing trajectory to an accelerated path with the ultimate target of full closure (Fig. 4A). This shift is achieved using a machine learning framework composed of three key components: a Deep Mapper, a model used to find an optimal healing path with a Linear Quadratic Regulator (LQR), and a Deep Reinforcement Learning (DRL) controller (Fig. 4B). The Deep Mapper comprises two main elements: an encoder and a decoder (Fig. 4C), both implemented via deep neural networks (DNNs). The encoder processes wound images to predict the healing trajectory by defining the wound state as probabilities across the four key healing stages: hemostasis, inflammation, proliferation, and maturation. Additionally, it calculates transition velocities between these stages (e.g., hemostasis to inflammation, inflammation to proliferation, proliferation to maturation). These velocities enable the construction of a linear dynamic model, ^18^ from which an optimal control law^37^ can be analytically derived using the LQR framework to minimize wound closure time. However, this LQR-derived optimal control is theoretical, representing adjustments to healing stages that minimize closure time but does not directly translate into actionable treatment parameters like electric field strength and drug dosage. To address this limitation, we implement a leader-follower strategy^38^, commonly used in robotic control^39^, that combines the decoder with a DRL controller. The decoder generates a projected image of the wound, reflecting its predicted appearance if optimally treated according to the LQR model. This projected image acts as the “leader,” representing the ideal healing outcome. In the final step (Fig. 4D), two DRL controllers, referred to as “followers,” are employed: one to determine the optimal fluoxetine (Flx) dosage and another to adjust the electric field (EF) intensity. These controllers aim to minimize the discrepancy between the actual wound image (post-treatment with Flx or E_F_) and the projected image, thereby ensuring that the healing process aligns closely with the optimized trajectory.^18^

**Fig. 4.**
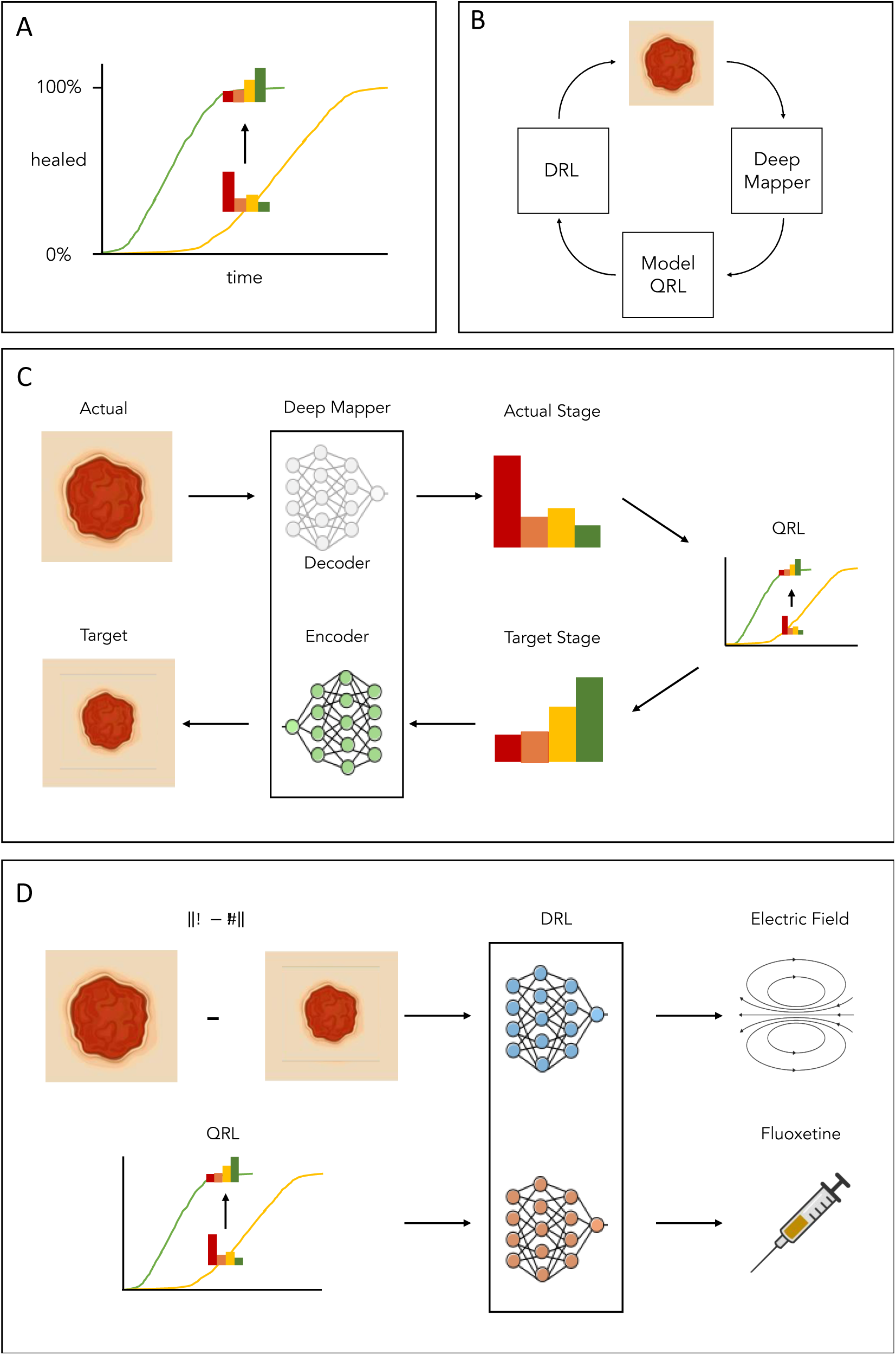
***Schematic Description of the AI Physician and its components.*** (A) Strategy for accelerating wound healing. (B) Control system components for wound healing acceleration Deep Mapper, Model with Quadratic Linear Regression (QRL), (C) functioning of deep mapper, (D) architecture for DRL to deliver optimal treatment.

### Porcine Wound Healing

We deployed the TheraHeal platform on a porcine excisional wound model to evaluate its effectiveness over a 22-day period ^40^, with the closed-loop therapy running during the first 7 days (Fig. 5A). The TheraHeal wearable attached to a 20 mm wound on the porcine model supported the healing process (Fig. 5B, S15-S17). Before operating the device on the porcine model, thorough biocompatibility tests confirmed its suitability (S10). Two wounds were treated with the device and four wounds served as control (Fig S18). The swine was able to freely move in its enclosure fitted with the devices and battery packs and underwent its normal routine (Fig. S19). Tissue samples provided histological insights into wound healing outcomes. Throughout the experiment, the camera module captured high-resolution wound images every two hours (Fig. S20), enabling the AI Physician to perform wound monitoring, diagnostics, and real-time decision-making for optimal therapy based on the wound evolving condition. A laptop served as the server, granting access to a graphical user interface through a web browser in clinical settings (Figs. 5C–F). This data is for one of the treated wounds (W6, S21). The other treated wound had a partial device failure at day 4 caused by the swine showering to cool itself off (W2, Fig. S22). This personalized approach enabled stage-specific interventions during the 7-day treatment period. The GUI displayed wound images (Fig S23), predicted wound stages, and healing rates compared to the control wound in the model, highlighting whether the wound progressed faster or slower (Figs. 5C, 5D). It also tracked device performance metrics, such as E_F_ treatment data (Fig. 5E) and drug dosage (Figs. 5F, S21). Camera images at day 17 indicate faster closure of treated wound (Fig. S 24) and tissue analysis confirms that bioelectronic delivery of Flx is effective (Fig. S25), and there is no systemic accumulation reducing the chances of off target effects (Fig. S26). In this care regimen, the TheraHeal platform applied EF therapy until the treated wound’s probability of inflammation dropped below 40%, signaling progression toward the optimal healing trajectory (Fig. 5F). At this stage, TheraHeal initiated Flx delivery, with the dosage determined by the AI Physician (Fig. 5F). We chose this regime for the ability of EF to reduce inflammation^41^ and from our data that indicates that Flx application is more beneficial later on during healing^23, 24^. In a clinical or remote care setting, the GUI allows the physicians to intervene manually, fine-tuning treatments as needed (Fig. S23). By day 22, during the late phase of wound healing, histo-morphological analyses evaluated the regenerated tissue to determine wound resolution. The wounds fully resurfaced, achieving 100% re-epithelialization (Fig. 5G, Fig. S27). Measurements of epidermal thickness and the number of Rete pegs (protrusions from the epithelium to the dermis, which indicate epidermal maturity) indicated further differentiation and maturation of the repaired tissue for both treated wounds compared to control (Fig. 5H, Fig. S28). The device-treated wound W6 had a statistically significant increase in epithelial thickness (Fig. 5H), while the combined wound data W2 and W6 had trending higher epithelial thickness (Fig. S 29). The difference between the successfully treated wound W6 and the wound whose intervention was interrupted W2 is not surprising. This difference further confirms the importance of the holistic approach combining TheraHeal wearable and the AI Physician.

**Fig. 5.**
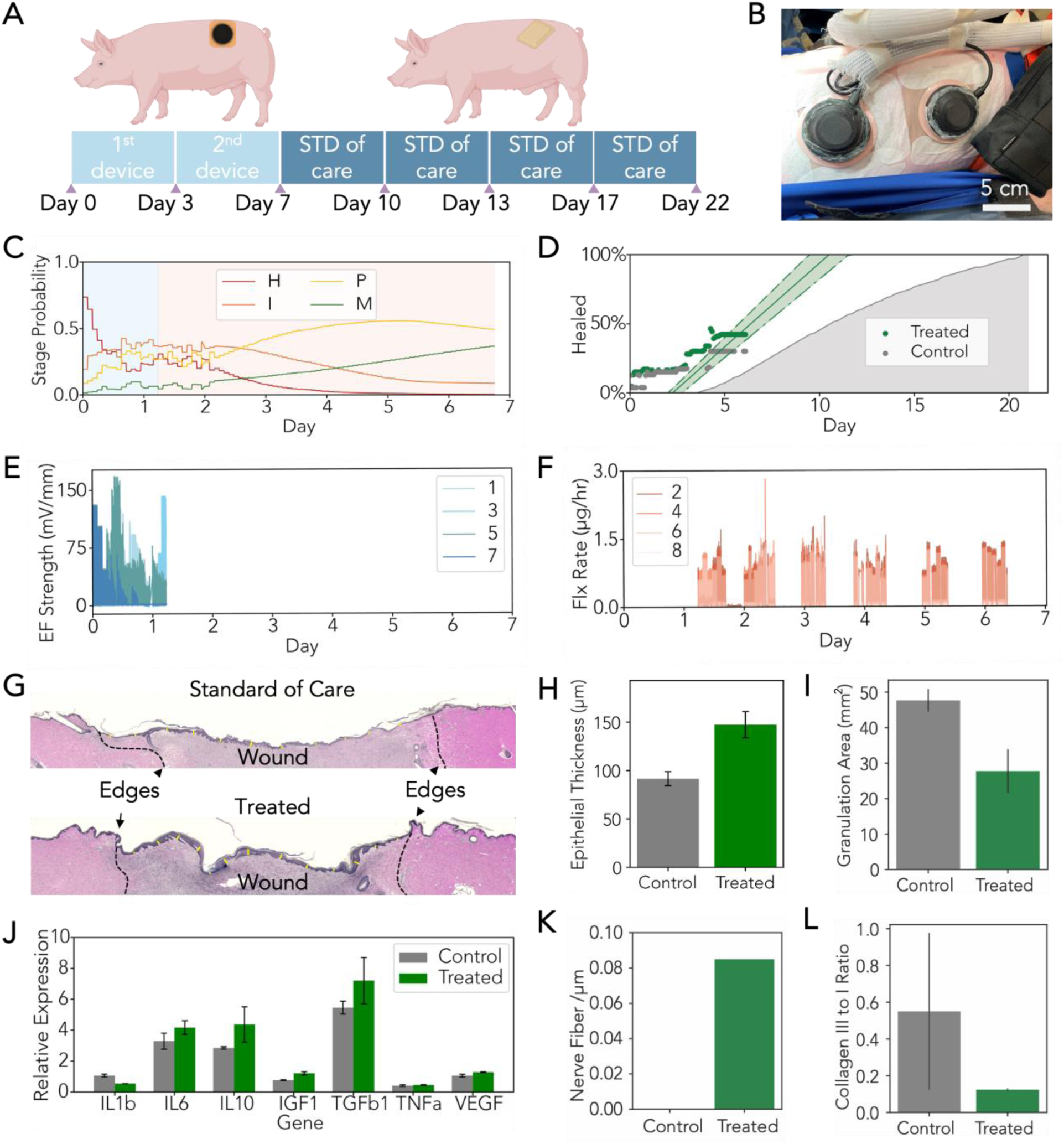
***In Vivo study in porcine model for 22 days.* (A) In vivo experiment schedule and setup.** (B) photo showing the device was mounted on 20 mm wound of the porcine model. (C) wound stage probability of a treated wound, interpreted by Deep mapper (D) Wound healing progress from control and treated wound, interpreted by Deep mapper. (E) Electric field strength from individual channels of a bioelectronic device. (F) Fluoxetine delivery rate from individual channels of a bioelectronic device. (G) Representative images from the day 22 wounds treated with standard of care (control) or the device. (H) Epidermal thickness was measured on multiple sites across the neo-epithelium covering the two control wounds and one treated wound W6(n_control_=30, n_treated_=16, p<0.01) (I) Granulation tissue was identified by difference in histological staining and outlined and area measured. (n= 2, p=0.03) (J) RT-PCR analysis of gene expression from healed wound tissue collected on day 22. Expression of anti-inflammatory IL10 and pro-preparative TGFB1 are elevated in the device treated wounds relative to control standard of care (n=2, IL1b p = 0.10; IL6 p = 0.33; IL10 p = 0.41; IGF1 p = 0.13; TGFb1 p = 0.44; TNF p = 0.57; VEGF p = 0.21). Genes associated with inflammation: IL1B, IL6, and TNFa are reduced in all healed wounds by day 22 (K) Nerve fibers identified by immunohistochemical tagging with antibody to PGP9.5, a protein expressed in peripheral nervous system axons, and imaged using confocal microscopy (Fig. S29) and linear structures within the epidermis tallied. (L) Ratio of collagen type III/I, derived from picrosirius red staining under polarized light from healed wounds at day 22 Fig. S29. (average +/-STDEV, * p<0.05, n=2 wounds)

Additionally, the wound bed granulation tissue area decreased by 41.8% in device-treated wounds compared to those receiving standard care, indicating faster progression into the regeneration phase and improved resolution of granulation tissue (Fig. 5I, S27). Gene expression analysis using Qiagen RT2 qPCR Primers for pigs found that expression of anti-inflammatory IL10 and pro-preparative TGFB1 are elevated in the device treated wounds relative to control standard of care. Genes associated with inflammation: IL1B2, IL6, and TNFa are reduced in all healed wounds by day 22 indicating faster progress of healing with treatment (Fig. 5J, Supplementary Methods or gene expression and qPCR). Immunohistochemical (IHC) analysis of the numbers of nerve fibers in the epidermis of the imaged area captures enhanced innervation after combination treatment^42^ (Fig. 5K and Fig. S30), less angiogenesis (Fig. S30), and a lower ratio of pro-inflammatory macrophages (M1) vs the pro-relatively macrophages (M2) (Fig. S31). During normal wound healing, fibroblasts deposit type III collagen to the wound site first, and later the matrix is replaced by type I collagen^43^. Therefore, the ratio between the type III to type I collagen points to progression of healing^44^. Here we used Picrosirius Red staining under polarized light to quantify the two types of collagens (Fig. 5L and Fig. S32). The decreased ratio in the device treated wounds indicates a more mature collagen structure^44^.

## Conclusions

TheraHeal is an AI-powered theranostic wound healing platform that enables continuous monitoring, diagnostics, and personalized on-demand wound care. This platform integrates AI image recognition to analyze simple images from an onboard camera, providing real-time wound diagnostics, with bioelectronic actuators that deliver personalized therapies, including electric fields and drugs. TheraHeal identifies wound stages and prescribes ad hoc therapies to guide each wound toward an optimal healing trajectory. TheraHeal continuously reassesses the wound and dynamically adjusts the therapy to match the evolving healing process. Physicians can monitor progress remotely via a graphical user interface, allowing for real-time assessment and intervention when needed. In a clinically tractable large animal model, TheraHeal accelerated healing with a combined electric field and fluoxetine therapy, showing improved epidermal thickness, reduced granulation tissue, and improved collagen I/III ratios. This portable, wireless platform makes physician-guided wound therapy accessible to patients with limited healthcare access, including those in remote areas or with limited mobility. Current limitations and future improvements include a flexible platform deployable on larger and irregular wounds. While tested on a healthy large animal model where physiological healing is already optimized, TheraHeal holds promise to improve healing in wounds where healing is impaired, such as chronic ulcers or diabetic wounds.

## Methods

### TheraHeal wearable Camera module fabrication

The imaging subsystem consists of an Arducam camera module with UC-665 ribbon cable, a plano-convex lens, and a custom LED PCB ring which are all housed within a 3D-printed enclosure. The enclosure components (bell top, lens holder, camera holder, LED PCB holder, and Bioelectronic Actuator holder) were fabricated using polylactic acid (PLA) on a Prusa i3 3D printer. Each holder features alignment interfaces that enable stacking of components and their respective housings, as depicted in the CAD model of the wound healing device. Prior to assembly, the LED PCB ring is designed in KiCad and arrives fully assembled from Sierra Circuits. M2 metal threaded inserts are also heat-set into 3D-printed camera holder. The Arducam module is then secured to the camera holder using two 6mm nylon M2 screws, and the UC-665 ribbon cable was inserted into the camera board.

Device assembly was performed under aseptic conditions in a Class II biosafety cabinet. All components underwent sterilization prior to assembly. Micro-HDMI cables were sterilized using a two-step protocol: thorough surface cleaning with 70% isopropyl alcohol (IPA) followed by UV irradiation for 15 min per side. Sterilized cables were stored in a stainless-steel container within the biosafety cabinet. Non-printed components were surface-sterilized with 70% IPA and UV-irradiated (15 min per side), while 3D-printed components were immersed in a 75:25 mixture of 99% IPA and deionized water prior to the UV exposure. The plano-convex lens was immersed in 70% IPA in a sterile polystyrene petri dish, with its curved surface oriented downward. Excess IPA was removed using non-woven wipes and silicon-tipped tweezers, followed by visual inspection to verify the absence of surface contamination.

Assembly proceeded sequentially under aseptic conditions. The LED PCB was positioned in its 3D-printed holder, with the micro-HDMI connector aligned with the access opening. The camera’s ribbon cable was also connected to the LED PCB, followed by placement of the cleaned plano-convex lens in its 3D-printed holder. The lens assembly, comprised of the lens and it’s corresponding holder, was then stacked onto the LED PCB assembly. The camera assembly was mounted above the lens stack, and a sterile micro-HDMI cable was connected to the LED PCB port. The assembly was completed by positioning the bell-top enclosure and securing it to the LED PCB holder using thermoplastic adhesive applied to the holder’s interfacing lip. The imaging subsystem is later combined with the bioelectronic actuator, its 3D printed holder, and skin barrier to complete the integrated device.

### TheraHeal Wearable PDMS Body Fabrication

The fabrication of the PDMS device involves molding three distinct components: the reservoir mold, the notch mold, and the channel piece. These molds are created using 3D printing and CNC milling techniques to achieve precision and consistency. Each of the molds is printed on a Formlabs 3B printer, using Model V3 resin with a layer thickness of 50 microns. To ensure optical clarity for the final device, a separate mold piece is fabricated from CNC-milled acrylic disks with a diameter of 19.5 mm. This step is critical for minimizing light scattering and optimizing the optical performance of the device.

The PDMS (Sylgard 184) mixture is prepared by combining the base and cross-linker in a 10:1 ratio. After thorough mixing, the mixture is degassed to remove trapped air, which helps prevent defects during molding. The prepared PDMS is then used to fill the reservoir and notch molds. The molds are carefully leveled and degassed again to ensure uniformity. Subsequently, they are cured at 60°C for 48 hours to ensure complete cross-linking and structural integrity. Once curing is complete, the PDMS components are extracted and undergo a cleaning process to remove any residual impurities. The components are sonicated in isopropanol (IPA), followed by thorough rinsing with water. Finally, the cleaned parts are oven-dried to prepare them for subsequent assembly.

### Design of PCB

A customized PCB is designed and send to a vendor (PCB way) to manufacture. The PCB is ring-shaped, with an outer diameter of 43 mm and an inner diameter of 20 mm. The channels consist of eight plated through holes (PTHs), each 1.75 mm in diameter, arranged in a circular pattern at a radial distance of 19 mm from the center. These PTHs allow for mechanical and electrical integration with the PDMS ion pump piece beneath the PCB. The center reference channel is a 0.7 mm diameter PTH connected to the electrical ground of the PCB.

The actuator PCB is an 8-channel wireless device for electrical actuation and sensing, with an additional center channel serving as the electrical ground (reference). It uses a WiFi-capable, dual-core microcontroller unit (MCU), the ESP32-PICO-D4, to perform low-level processing and communication tasks between various onboard hardware components. A custom device firmware is loaded onto the flash memory of the MCU for this purpose. Each channel can provide an applied voltage of up to 3.3 V and an output current of up to 330 µA, depending on the load resistance at the channel. The applied voltage is controlled using an onboard 8-channel DAC53608 digital-to-analog converter (DAC). Voltages are measured by two onboard 8-channel ADS7828 analog-to-digital converters (ADCs). Op-amp voltage buffer amplifiers are used at the input of the ADCs to provide high input impedance for accurate current and voltage measurements. Current is determined by sensing the voltage across a high-precision (0.1%) 10 kΩ resistor in series with each channel. The onboard MCU uses the inter-integrated circuit (I²C) interface to send commands to the DAC and acquire data from the ADCs. Measured data is stored locally on a microSD card and wirelessly transmitted to a remote laptop via a 3D metal antenna. The MCU communicates with the microSD card using a serial peripheral interface (SPI) and transmits data via WiFi using the TCP/IP protocol. Upon powering up, the PCB automatically connects to a local WiFi network set up with a commercial router. The remote laptop, also connected to the same network, sends high-level commands to the PCB for user-specified control, closed-loop operation, and real-time data acquisition. When the PCB receives a high-level command from the laptop, the firmware breaks it down into a sequence of low-level tasks for the MCU to execute.

### PCB Integration and Waterproof Coating on Actuators

Silver wires were inserted into each reservoir of the PDMS device to serve as working electrodes. These wires were then connected to the PCB. To ensure a proper seal, PDMS seals were applied at each insertion point and cured thoroughly. To assemble the device, custom-made aluminum clamps were used to hold the two parts of the device together during the plasma bonding process. The device parts were exposed to plasma for 10 sec to activate the surfaces, after which the clamps were immediately bolted to secure the assembly. The entire device was subsequently heated at 60°C for 30 min to facilitate bonding. The quality of the bonding was verified by injecting air into each reservoir and inspecting for any leaks. To protect the optical window of the device, adhesive tape was applied before the parylene coating process. The device was coated with a 2 g layer of parylene over a 3-hour period, ensuring an even and durable protective layer. Once the PCB was attached, each wire was carefully soldered to the corresponding plate through holes on the board. A chlorinated silver wire was inserted into the central channel of the device to function as a counter electrode. Finally, a second layer of parylene coating was applied to provide additional protection and ensure the durability of the entire assembly.

### Assembly of Actuator

Capillaries (800 µm ID, CT95-03: TLC Spotting Capillary Tubes) were filled with NaOH for cleaning and subsequently conditioned using deionized water (MQ Water, 18.2 Ω), A174 silane coupling agent, and ethanol. This conditioning process was performed using a syringe pump to ensure consistent flow rates and thorough treatment of the inner surfaces. Following this, the capillaries were thoroughly dried to eliminate any residual solvents. The dried capillaries were then filled with the hydrogel precursor solution. Ultraviolet (UV) curing was applied to polymerize the hydrogel within the capillaries, using a UV intensity of 7 mW/cm² for an adequate curing time. The cured hydrogel-filled capillaries were cut into segments of 5 mm each. These segments were subsequently soaked in either Flx or Steinberg solution, depending on the intended application of each channel. For device preparation, reservoirs and channels were filled with the corresponding solution, Flx or Steinberg solution, based on whether the channel was designated for Flx or EF delivery. Capillary segments were inserted into each channel using a biopsy punch to create precise openings for integration. To ensure the structural integrity of the device, a biocompatible cap was 3D-printed and used to seal each channel effectively. This cap was specifically designed to prevent leakage and maintain a sterile environment.

Control software was loaded onto a custom PCB, allowing for accurate monitoring and regulation of the functions of the device. A thorough functionality test was performed to validate each aspect of the system, ensuring that it operated correctly before integration into the camera harness for experimental use.

The device and enclosure were then glued to a skin barrier (Hollister New Image™ Flat CeraPlus™ Skin Barrier – Tape,11204) which come with adhesive that can mount the device to the wound.

### Pig acclimation and wounding surgery

The Yorkshire-Landrace-Duroc cross breed pigs (30-55 Kg, young females) were ordered from the UCD Swine Research Center (Davis, CA). The procedure for swine care and the wounding surgery ^45^ were derived from the guidelines of the Association for Assessment and Accreditation of Laboratory Animal Care and the NIH Guide for the Care and Use of Laboratory Animals, reviewed, and approved by the UC Davis Campus Veterinary Services, and the UC Davis Institutional Animal Care and Use Committee. The animals were acclimated for 7-10 days, trained for saliva collection (saliva induced with fresh apple slices, and 0.5-1mL collected with 4 large cotton swabs swiped across the mouth cavity in the morning before 12:30pm for cortisol measurement by HPLC), harness use (Kruz Original Heavy-Duty No Pull Dog Harness, medium to extra-large sizes), and human contact with positive reinforcement (food treats), and fasted for 15 hours the night before the surgery. Anesthesia induction was with Telazol (5.5 mg/kg, intramuscular injection), followed by masked inhalation of Isoflurane (1-5% to effect). Then the pig was intubated, and body temperature, heart rate and respiratory rate were monitored every 15 minutes. The pig blood samples were collected from the ear vein (up to 10mL per collection) for plasma and serum isolation. The dorsal skin was shaved, hair-removed with depilation cream, and prepared with 2% chlorhexidine scrubs and isopropyl alcohol rinses. Prophylactic antibiotic (Cefazolin, 30-35 mg/Kg via IV) was injected during the wounding surgery.

Six circular wounds (20mm in diameter, at least 50 mm apart to fit the device and skin barrier) were created with a scalpel blade and scissors down to the desired depth (partial thickness wounds, 6mm deep) on the paravertebral area of a pig. After compression hemostasis, wound images were captured with a cell phone camera (iPhone 7, Apple) held at a constant distance of 10 cm above the wound, with a wound scale (Medline NE1 Wound Assessment Tool). The wounds were rinsed with a 0.001% cefazolin and then received standard care or the sterile devices (device-treated). The standard care wounds were covered with Conformant 2 Wound Veil (Smith & Nephew, Watford, England, UK), a hydrophilic polyurethane foam dressing, Optifoam (Medline, Illinois, USA), and a transparent dressing, Tegaderm (3M, Minnesota, USA). The devices were pre-tested for Wi-Fi connection and applied to the wounds. The skin barrier adhesive dressing on the devices was further secured with MedFix Dressing Retention Tape (Medline). The entire dorsum of the animal with the wounded area was protected with foam sheets (3M, Reston self-adhering foam pad) in a spandex tube (Showpro Lamb tube) with Velcro openings on the back to access the wounds. The devices were connected to the power bank (Voltaic Systems V75) and the Raspberry Pi (model 4B) controller, carried in 2 pouches (Tacticool Molle) attached to the shoulder harness on the pig. The exposed wires between the devices and the pouches were covered with cord protector tubing to prevent chewing by the animal. Post-operative analgesic (buprenorphine, 0.005-0.1 mg/kg intramuscular injection before extubation; fentanyl patch, 1-5 ug/kg/hr for 72 hours) was given at the end of the surgery, and the pig was monitored for signs of hyperthermia after recovery.

### Device actuation

The devices were connected to a designated wifi network (Fig. S15-16), set up in the vivarium. The researchers operate the device through wireless command from the laptop computer.

Camera control and image acquisition were implemented using libcamera on Linux, which enables command-line configuration of the Arducam module. The imaging protocol consists of three scripts: a preview script using continuous image capture for real-time monitoring, an acquisition script with various parameters for use during experiments, and a validation script that performs automated capture of six sequential images in a 24-hour stress test cycle. The image acquisition parameters include a 70s initialization delay to ensure system stability and network connection, followed by 3s programmed delay between image captures. The focal parameter is varied from representative values 0 to 5 to capture z-stack images, with a value of 3 corresponding to the theoretical focal plane. Each Raspberry Pi unit is configured with a unique camera identifier corresponding to its designated wound site, and automated image transfer to remote storage is implemented using an IP address. The system includes an automated shutdown protocol for reduced power consumption post-acquisition during in vivo experiments.

The actuator automatically connects to Wi-Fi and enters standby mode. A software module, incorporating the Deepmapper and DRL algorithm, runs on a laptop, sending commands to the actuator. The module includes a user-friendly GUI that allows researchers to monitor and interact with the system. The software automatically stops the actuator when the target treatment is achieved, or the set duration expires.

To maintain consistency across experiments, the device and software operate for 22 hours daily, with a 2-hour window allocated for data transfer, battery replacement, and routine checks. The battery (Voltaic Systems V75), which can support 36 hours of operation, is typically replaced daily to ensure uninterrupted power. After each battery change, the software module is manually reset, and data are copied to the laptop’s hard drive. This process is repeated daily throughout the experiment.

### Post-operative care

The wounds and the devices were examined daily. The wound device and dressings were replaced every 3-4 days over the course of the experiments. At the end of observation on postoperative day 22, a blood sample was again collected, and the animal was euthanized. Wound tissue was excised with a 10 mm margin of surrounding intact skin, bisected along the axis of the device actuation and the channels reaching the targeted treatment concentration, formalin-fixed, paraffin-embedded, and sectioned to 5μm for H&E staining and immunohistochemistry (IHC).

### Wound epithelialization

Wound tissues were fixed in 4% paraformaldehyde for one week, embedded in paraffin blocks (Tissue Tek processing and embedding stations, Sakura Finetek, Torrance, California) and 5 um thick sections were affixed to glass slides. After drying, the sections were stained to visualize neo-epithelium, collagen deposition, neurons, or macrophages. Tissue sections were stained using a standard H&E protocol to assess re-epithelialization as previously described ^45^. Stained sections were imaged using a BZ-9000 inverted microscope, and the images were scored in the BZ-II Analyzer software (Keyence, Osaka, Japan). The wound edge was identified by the absence of subcutaneous fat and muscle and by the visible demarcation between the granulation tissue and mature dermal collagen. Re-epithelialization was measured along the basal keratinocytes, and the percentage was calculated by the length of the neo-epithelium from both edges and the total length of the wound.

### Picrosirius Red staining and collagen ratio

Five-micron wound sections were stained with Picrosirius Red solution (American MasterTech/StatLab, McKinney, TX) for 1 hour, destained with acidic alcohol according to the manufacturer’s instruction, and imaged under polarized light ^46^ on a BioRevo BZ-9000 inverted microscope (Keyence, Japan) with the BZ-Viewer and BZ-Analyzer software (Keyence, Japan). Color extraction of type I collagen (red to orange color) and type III collagen (green to yellow color) was performed with the “hybrid cell-count” function in the BZ-Analyzer. The area of the two types of collagen in the wound bed (identified by H&E staining in a sequential section) was quantified in each section for the collagen III to I ratios. High ratio of collagen III indicates newly regenerated granulation tissue, whereas high ratio of collagen I suggests well re-organized, mature dermal tissue.

### Cortisol extraction from saliva and Chromatographic Analysis

Pig saliva was expressed from the cotton swabs and centrifuged to remove particulate matter. The clarified saliva was divided into aliquots and stored at-80°C until analysis. Samples were extracted according to a method described by Hofreiter et al. (PMID: 7147286), modified for measurement of cortisol in saliva. Briefly, 0.300 mL saliva was shaken for 3 minutes with 1.000 mL dichloromethane. The aqueous phase was removed, and the dichloromethane layer was washed by shaking for 1 minute with 0.100 mL 0.1 N NaOH followed by 0.100 mL water. After each step, the samples were centrifuged to induce phase separation, and the aqueous phase was removed by aspiration. The dichloromethane layer was transferred to a clean tube and evaporated at 40°C under a gentle stream of nitrogen. The residue was reconstituted in 100 uL of 50:50 methanol: water, and 10 uL was injected for analysis.

Measurements were performed using reverse phase HPLC with UV absorbance detection. The system was an Antec Neurotransmitters Analyzer coupled to a Knauer BlueShadow 40D detector. Separation was performed on a Waters Acquity UPLC BEH C18 column (3.0 mm x 100 mm, 1.7 um) at a flow rate of 0.450 mL per minute and a temperature of 35°C. The mobile phase consisted of 28:72 acetonitrile: water, and column effluent was monitored at 242 nm.

### Extraction of Fluoxetine and Norfluoxetine from Pig Plasma

Pig plasma samples were purified prior to analysis using C18 cartridge solid-phase extraction (SPE). 100 mg C18 cartridges were packed into glass SPE tubes with glass fiber frits (Macherey-Nagel, Duren, Germany). The sorbent was taken from Sep-Pak Plus 360 mg C18 cartridges (Waters, Ireland) which were broken open. Plasma extraction was carried out as follows: cartridges were placed onto a vacuum manifold and conditioned with two tube volumes of methanol and one tube volume of water. Pig plasma (0.500 mL) was spiked with 50.0 uL internal standard solution (40.0 ug/mL fluvoxamine in methanol), diluted with 0.500 mL H_2_O, and this was passed through the conditioned SPE cartridges. Next, cartridges were rinsed with one tube volume H2O followed by one tube volume 50:50 methanol: H_2_O. Fluoxetine and norfluoxetine were eluted from the cartridges in 0.500 mL methanol containing 0.5% (v/v) formic acid, the eluate was made up to 1.000 mL by addition of 0.500 mL H_2_O, and 10 uL of this was injected for analysis.

### Immunohistochemistry Staining of Pig Skin

Formalin-fixed, paraffin-embedded (FFPE) skin sections from day 23 were used for immunohistochemistry (IHC) staining to visualize macrophages, neurons, and angiogenesis.

Macrophages were labeled with antibodies against iNOS (M1; ThermoFisher, Catalog # PA1-036), arginase 1 (M2; ThermoFisher, Catalog # PA5-18392), and a pan-macrophage marker (BA4D5; Bio-Rad, Catalog # MCA2317GA). Multicolor labeling was achieved using donkey anti-goat Alexa Fluor 568 (Invitrogen, Catalog # A-11057), donkey anti-rabbit Alexa Fluor 647 (Invitrogen, Catalog # A-31573), and donkey anti-mouse Alexa Fluor 488 (Invitrogen, Catalog # R-37114). Neurons were identified in consecutive skin sections of 30 µm thickness using the PGP9.5 antibody (ThermoFisher, Catalog # 480012) and donkey anti-mouse Alexa Fluor™ 488 (Invitrogen, Catalog # R37114) as the secondary antibody. Angiogenesis was assessed with the CD31 (PECAM-1) antibody (Cell Signaling, Catalog # 77699S), followed by donkey anti-rabbit Alexa Fluor 647 (Invitrogen, Catalog # A-31573). All slides were mounted with VECTASHIELD® Antifade Mounting Media with DAPI (Vector Laboratories, Catalog # H-1200-10) for nuclei visualization. Imaging was performed with a Zeiss LSM900 Confocal Laser Scanning Microscope at a resolution of 3 pixels per µm. For each wound sample, three stitched images of the wounded region near the healed epidermis were collected using a 40× oil immersion lens to minimize tissue variation. Quantification of macrophages, neurons, and blood vessels was performed by visualizing the staining in the confocal images, and analysis was carried out accordingly.

### Extraction of Serotonin from Pig Plasma

Prior to analysis, pig plasma samples were depleted of protein as follows: pig plasma (0.100 mL) was spiked with 5.00 uL antioxidant solution (1.3% (m/v) ascorbic acid and 2 mM EDTA in H_2_O) and 20.00 uL internal standard solution (300.0 uM N-methylserotonin in H_2_O). Next, 5.64 uL of 70% perchloric acid solution (Thermo Scientific, Waltham, Massachusetts) was added and the samples were vortexed for 30 seconds. The samples were centrifuged, and 10 uL of the supernatant was injected for analysis.

### Fluoxetine concentration measurement from wound tissue

Pig wound tissues (20.0 to 30.0 mg) were cryo-pulverized over liquid nitrogen, resuspended in 0.250 mL methanol containing 0.5% (v/v) formic acid, and sonicated for 15 minutes in a sonicating bath. The tissue homogenates were centrifuged, the supernatants were made up to 0.500 mL by addition of 0.250 mL water, and 10 uL of the finished extract was injected for analysis.

Measurements were performed using reverse phase HPLC with UV absorbance detection. Separation was performed on an Acquity UPLC BEH C18 column (3.0 mm ID x 100 mm L, 1.7 um particles) at a flow rate of 0.450 mL per minute and a column temperature of 35C. The mobile phase consisted of 35:65 acetonitrile: 45 mM phosphate buffer pH=6.0, and column effluent was monitored at 230 nm.

### Chromatographic Analysis of Fluoxetine, Norfluoxetine, and Serotonin

Measurements were performed using reverse-phase HPLC with UV absorbance detection. The system was an Antec Neurotransmitters Analyzer coupled to a Knauer BlueShadow 40D detector. Separation was performed on a Waters Acquity UPLC BEH C18 column (3.0 mm x 100 mm, 1.7 um) at a flow rate of 0.450 mL per minute. For fluoxetine and norfluoxetine analysis, the mobile phase consisted of 35:65 acetonitrile: 45 mM phosphate buffer pH = 6.0, separation was performed at 35°C, and the column effluent was monitored at 230 nm. For serotonin analysis the mobile phase consisted of 4:96 acetonitrile: 50 mM citrate buffer pH = 4.3, separation was carried out at 37°C, and the column effluent was monitored at 275 nm.

### Gene expression and qPCR

Wound tissue was preserved in Invitrogen RNALater Stabilization Solution (Fisher Scientific) at 4 degree for 4 days and frozen at-80 degree according to the manufacturer’s instructions. The frozen tissue was trimmed with a 5mm biopsy punch from the wound edge (50 mg tissue), minced to 1mm pieces, and homogenized with Tissue Tearor (BioSpec cat# 98537004) in 300 µl Buffer RLT containing β-mercaptoethanol (Qiagen RNeasy Fibrous Tissue Mini Kit) on ice for 2 minutes. The mRNA was extracted with Qiagen RNeasy Fibrous Tissue Mini Kit, and cleaned with DNase I according to the manufacturer’s instructions. One μg of RNA was reverse transcribed to cDNA using the Quantitect Reverse Transcription Kit (Qiagen). The following Qiagen RT2 qPCR Primer for pigs were used: IL1B2 (PPS00015A-200), IL6 (PPS00991A-200), IL10 (PPS00445B-200), IGF1 (PPS00801A-200), TGFB1 (PPS00418A-200), TNF (PPS00426A-200), VEGFA (PPS00495A-200), NOS2 (PPS00221A-200), ARG1 (PPS00454A-200), DCX (PPS14382A-200), MAP2 (PPS02696A-200), COL1A1 (PPS72004A-200), COL3A1 (PPS02256A-200), RPL19 (PPS00333A-200), and GAPDH (PPS00192A-200). For qPCR analysis, the Quantitect Sybr Green master mix was used (Qiagen) on a 384-well platform (Bio-Rad CFX384 Real-Time System, C1000 Touch Thermal Cycler). Each sample was run in duplicate and mRNA levels of target genes were normalized to the average levels of housekeeping genes GAPDH and RPL19. Relative expression of the target gene was calculated via ΔΔCt against the average Ct of the housekeeping genes and the average Ct of day 0 skin from each animal. The fold change dataset was analyzed by the Grubbs Tests to remove outliers, and by the t-test to determine statistical significance.

## Data Availability

All data produced in the present study are available upon reasonable request to the authors

## Acknowledgments

AI assisted technologies (ChatGPT) were used to revise the draft manuscript for current grammar and improved readability of the content written by the authors.

## Author contributions

Conceptualization: MR, RI, MG, MT, MZ

Methodology: MR, RI, MG, MT, HY, AS, MZ, HL, FL, WH, NA

Investigation: HL, HS, FL, WH, NA, PB, AG, KS, KZ, CR, MT, MK, GK, TN, SF, CF, KD, AB, SC, AE, KL

Visualization: HL, GL, SK,

Funding acquisition: MR, RI, MG, MT, MZ

Project administration: MR, EA

Supervision: MR, RI, MG, MT, MZ, AS

Writing – original draft: HL, GL, SK, HS, MR, MT, MT, MG, FL, RI

Writing – review & editing: MR, RI, MG, MT, MZ, AS

## Data and materials availability

All data, code, and materials used in the analysis are presented in the manuscript or the supporting information.

## Supplementary Materials

Supplementary Textf

Figs. S1 to S32

Tables S1 to S2

Data S1 to S2

