## supplemental file for "Real-time diagnostics and personalized wound therapy powered by AI and bioelectronics"

**The PDF file includes:**

Supplementary Text
Figs. S1 to S32
Tables S1 to S2
References

**Other Supplementary Materials for this manuscript include the following:**

Data S1 to S2

**Supplementary Text**

Code for closed-loop modules

Codes were shared open source on Github with the following link:
Deep mapper for wound stage analysis
https://github.com/Fan-Lu/DeepMapper

Deep reinforced learning decision maker
https://github.com/Fan-Lu/RL4Wound

Design of Actuator PCB

The actuator PCB is an 8-channel wireless device designed for electrical actuation and sensing, with an additional center channel serving as the electrical ground (reference). It features a ring-shaped design with an outer diameter of 43 mm and an inner diameter of 20 mm. The eight channels are implemented as plated through holes (PTHs), each 1.75 mm in diameter, arranged in a circular pattern at a radial distance of 19 mm from the center. These PTHs facilitate mechanical and electrical integration with the PDMS ion pump piece beneath the PCB. The center reference channel consists of a 0.7 mm diameter PTH connected to the PCB's electrical ground. The PCB is equipped with a WiFi-capable, dual-core microcontroller unit (MCU), the ESP32-PICO-D4, running custom firmware to handle communication with onboard hardware components. Actuation is performed by current-source digital-to-analog converters (DACs), and sensing is carried out by analog-to-digital converters (ADCs), both operating at a supply voltage of 4.7 V. Each channel can provide an applied voltage of up to 4.7 V and an output current of up to 250 µA, depending on the load resistance. Two 4-channel DAC63004 current-source DACs control the applied voltage, while two 8-channel ADS7828 ADCs perform the voltage measurements. Operational amplifiers (op-amps) configured as voltage buffers are placed at the ADC inputs to provide high input impedance. Current is determined by measuring the voltage across a high-precision (0.1%) 10 kΩ resistor at each channel. The MCU communicates with the DACs and ADCs using an inter-integrated circuit ($I^2C$) interface. Measured data is stored locally on a microSD card and wirelessly transmitted to a remote laptop via a 3D metal antenna. The MCU communicates with the microSD card using a serial peripheral interface (SPI) and transmits data via WiFi using the TCP/IP protocol. The PCB is powered by a 5 V output from a power bank, regulated through two separate low-dropout (LDO) regulators to generate 4.7 V and 3.3 V outputs. The 4.7 V supply powers the DACs, ADCs, and op-amps, while the 3.3 V supply powers the MCU and the microSD card. To facilitate $I^2C$ communication between the MCU and the DACs/ADCs, a 3.3 V to 4.7 V level translator chip handles logic level differences.
Upon startup, the PCB automatically connects to a local WiFi network established by a commercial router. The remote laptop, connected to the same network, sends high-level commands to the PCB for user-specified control, closed-loop operation, and real-time data acquisition. The firmware then translates these high-level commands into low-level tasks for the MCU to execute.

To ensure efficient multitasking, the ESP32-PICO-D4 MCU is programmed to divide tasks between its two cores: Core 0 adjusts the output voltages of the DACs to achieve specified target currents and samples data from the ADCs, while Core 1 handles TCP/IP commands for WiFi communication with the remote laptop and manages microSD card read/write operations.

PID Control of current for actuation

a proportional-integral-derivative (PID) controller with an anti-windup function runs on all 8 channels of each bioelectronic actuator to achieve and maintain the target current levels using The PID controller runs on a remote laptop and communicates wirelessly with the actuator PCB to facilitate feedback control. The control process begins by comparing the target current with the feedback current and gradually adjusting the applied voltage until the target current is attained at each channel. To maintain the target levels, the PID controller continuously adjusts (increases or decreases) the applied voltage based on the feedback current received wirelessly from the actuator PCB. Channel currents can vary due to the dynamic nature of contact or wound resistance, which is influenced in part by animal movement. Consequently, continuous feedback and real-time voltage adjustments are necessary to maintain the desired current levels. The anti-windup function enhances the system's responsiveness to changes in target current by clamping the integral term and PID output when saturation occurs. If the applied voltage reaches its maximum limit and the target current is still unattainable, voltage pulses (e.g., 3.3 V for 60 s, followed by 2 V for 5 s) are applied to generate high charging currents. Once the target current is achieved, these pulses are discontinued, and normal PID control resumes to maintain the target level.

EF Strength calculation.

We used two calculation methods for EF strength. One is directly simulated by finite element simulation (COMSOL) by loading the CAD geometry of the bioelectronic device, with a 5 mm thick layer as tissue with the conductivity of 0.17 S/m, which is an average of muscle layer at different directions[1]. Results are shown in Fig. 3E and Fig. S7(with different scale). The simulation also gives us the voltage distribution and the resistance of the wound, which is 31.1 kΩ. A nominal EF strength is calculated based on this resistance value, the current measured from potentiostats on board, and the distance of 8 mm between the working and ground port.

$$E = \frac{V}{d} = \frac{I \cdot R}{d}$$

Where:

$E$ is the nominal electric field strength (mV/mm),

$I$ is the current (uA), which is measured and recorded using the potentiostat on board.

$R$ is the resistance (kΩ), 31.1 kΩ.

$d$ is the distance (mm), which is 8 mm based on the geometry of the device.

The nominal value is used in plot EF over time (Fig. 5C)

Fluoxetine dose calculation

Fluoxetine was delivered using a bioelectronic ion pump, as described previously[2]. In this system, fluoxetine is protonated under slightly acidic conditions (pH ~6) to form the positively charged $Flx^+$ cation. The $Flx^+$ ions are then transported via electrophoresis, moving from the working electrode to the ground electrode along the direction of the electric field.

Since $Flx^+$ is the primary ion in the source reservoir, the amount of fluoxetine delivered is directly proportional to the total electrical charge (i.e., the number of electrons) passing through the circuit. To determine the delivery efficiency, we used HPLC to generate a calibration curve from a series of standard samples (Fig. S9). By integrating the current over time to calculate the total charge (Fig. S10), we quantified the efficiency of the devices (Table S1).

Biocompatibility test

Before conducting experiments on the porcine model, we evaluated the biocompatibility of our bioelectronic device components in collaboration with Da Vinci Biomed Research, Inc. This involved implanting PDMS and hydrogel components into a porcine model and observing immune responses. Six animals were included in the study, tested with three groups of different device configurations: PDMS with parylene coating, PDMS with parylene coating and empty glass capillary inserted, and a full set of PDMS with parylene coating and insert of capillary filled with ion-exchange hydrogel. These configurations mimic the interface of the bioelectronic device when in contact with a wound.

All six animals were successfully implanted with the devices in a full-thickness dermal wound defect model. Five animals survived the 30 ± 1-day study period as planned, with no health concerns attributable to the implants. At necropsy, no visible signs of infection or unusual inflammatory responses were observed in the wound beds. Blood tests revealed mostly normal results. Two exceptions were noted: Animal ID 1375 showed a high percentage of eosinophils, and Animal ID 1372 had immature neutrophils (absolute bands) on Day 0. These findings were isolated and did not impact the overall health of the animals. Histopathological analysis confirmed that the PDMS devices (Test Devices 1 and 2) were safe and well-tolerated at the endpoints (Day 29 or 30). Inflammation scores were minimal to mild for all devices, with occasional moderate scores for Test Device 2 at specific implant sites. Approximately half of the sites implanted with Test Device 2 showed detectable basophilic foreign material, likely residual hydrogel. Compared to the Control Device, reactivity scores were minimal or absent for Test Device 1 and slightly elevated for Test Device 2. No significant abnormalities were observed in downstream tissues aside from expected background lesions.

In conclusion, the PDMS device developed by UCSC demonstrated good biocompatibility and safety in a full-thickness dermal wound defect model over a 30-day period. The device did not cause unusual inflammatory responses or adverse effects in surrounding tissues. A detailed report is provided in the Supplementary Information file.

Flowcytometry for biocompatibility

During the treatment experiment, we also collected tissue samples from the control and treated wounds. The skin was mechanically and enzymatically digested to isolate single cells. Single cells were stained with antibody panels specific for myeloid cell populations, such as macrophages and neutrophils. There was no difference in total immune cells (CD45+), macrophages (CD45+2B2−BA4D5+), or neutrophils (CD45+2B2+) between control (standard of care) and control device, indicating that the device is compatible with skin and does not cause an immune response.

Topical fluoxetine wound treatment did not produce systemic effects in the pig

Pig plasma was collected at the endpoint of the experiment and analyzed for the presence fluoxetine and norfluoxetine, the primary active metabolite found in the blood of patients undergoing oral fluoxetine therapy. The therapeutic window for the combined concentration of fluoxetine + norfluoxetine in the blood of patients undergoing oral fluoxetine therapy is 120 to 500 ng/mL[3]. The limits of detection for our analysis were 8.1 ng/mL fluoxetine and 6.4 ng/mL norfluoxetine in pig plasma. As shown in **Fig. S26a,** neither analyte was detected.

Plasma serotonin was measured prior to wounding and at the experimental endpoint in order identify possible changes resulting from the topical fluoxetine wound treatment. Serotonin was analyzed in the plasma of 13 sex- and age-matched pigs from the UC Davis farm colony to establish the normal plasma serotonin values in this cohort of animals. As shown in **Fig. S26b** the serotonin concentration in plasma collected at baseline and following treatment using the experimental device did not differ from the normal plasma serotonin values for the cohort.

Systemic consequences of topical administration of fluoxetine to porcine skin wounds was probed by analysis of plasma levels of both fluoxetine and its metabolite norfluoxetine. Fluoxetine and norfluoxetine were not detected in the plasma collected at the experimental endpoint, indicating that systemic effects are unlikely to result from topical fluoxetine wound treatment. Plasma serotonin was measured prior to wounding and at the experimental endpoint in order to identify possible changes resulting from the topical fluoxetine wound treatment. In patients undergoing oral fluoxetine therapy for the treatment of major depressive disorder (MDD), plasma serotonin increased by ~60% at 10 hours following oral fluoxetine administration and had returned to baseline levels by 24 hours[4]. In another study of patients taking SSRIs for the treatment of MDD, daily plasma serotonin levels decreased by ~60% at the end of 8 weeks of daily oral SSRI treatment compared to baseline levels measured before starting treatment.[5] The serotonin concentration in plasma collected at baseline and following treatment using the experimental device did not differ from the normal plasma serotonin values for the cohort.

<u>Fluoxetine concentration measurement in tissue</u>
Pig wound tissue was collected following topical fluoxetine treatment using the experimental device. The final dose of fluoxetine was applied on day 6, and tissue was collected at the experimental endpoint on day 7 or 22. Tissues concentrations of fluoxetine were interrogated using HPLC-UV. On day 7 of healing, the tissue fluoxetine concentration was 5.98 ± 1.50 ng/mg tissue, and by day 22 of healing no residual fluoxetine was detectable in the wound tissue.

Animal experiments were conducted under UC Davis IACUC protocol #23353

### <u>**References**</u>


1. Gabriel C, Peyman A, Grant EH. Electrical conductivity of tissue at frequencies below 1 MHz. *Phys Med Biol* **54**, 4863-4878 (2009).

2. Li H, *et al.* Programmable Delivery of Fluoxetine via Wearable Bioelectronics for Wound Healing In Vivo. *Advanced Materials Technologies* **9**, 2301115 (2024).

3. Fluoxetine, Serum (test# 80228).). Mayo Clinic Laboratories (2024).

4. Blardi P, *et al.* Serotonin and Fluoxetine Levels in Plasma and Platelets After Fluoxetine Treatment in Depressive Patients. *Journal of Clinical Psychopharmacology* **22**, 131 (2002).

5. Holck A, *et al.* Plasma serotonin levels are associated with antidepressant response to SSRIs. *Journal of Affective Disorders* **250**, 65-70 (2019).

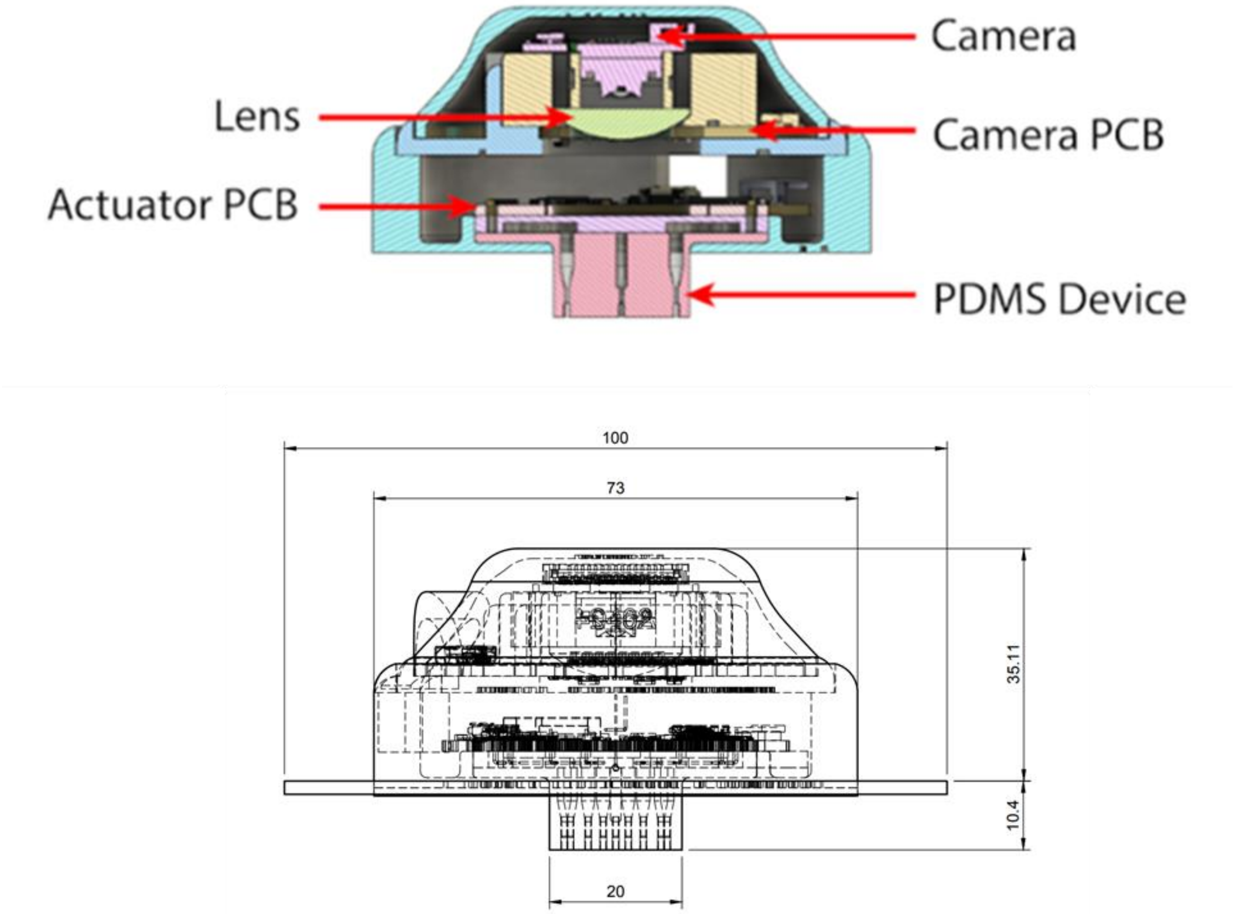


**Fig. S1**
Dimension of the TheraHeal wearable

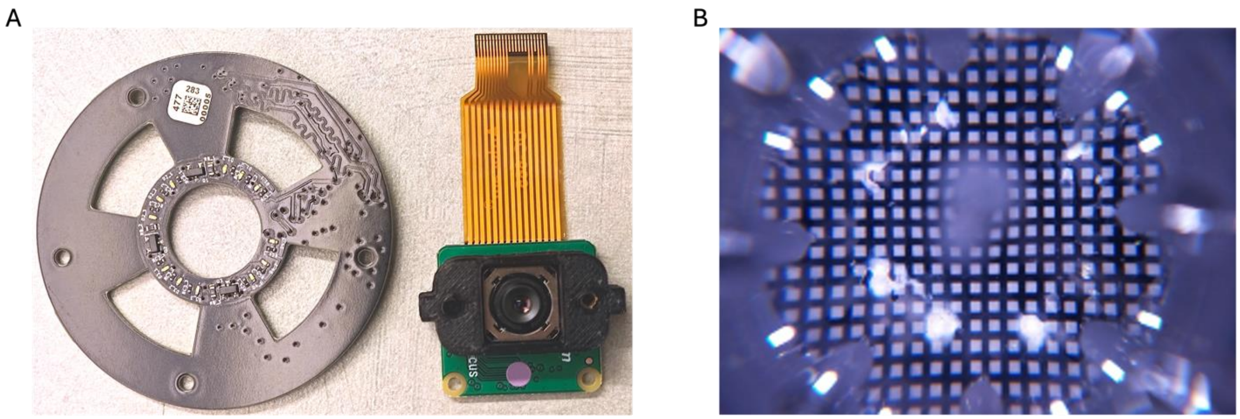


**Fig. S2.**

A. LED array (left) and camera (right) used in the final design of the device. B. Example of a calibration image. The calibration images are taken as part of the imaging Quality Control process to quantify the distortions caused by the PDMS device.

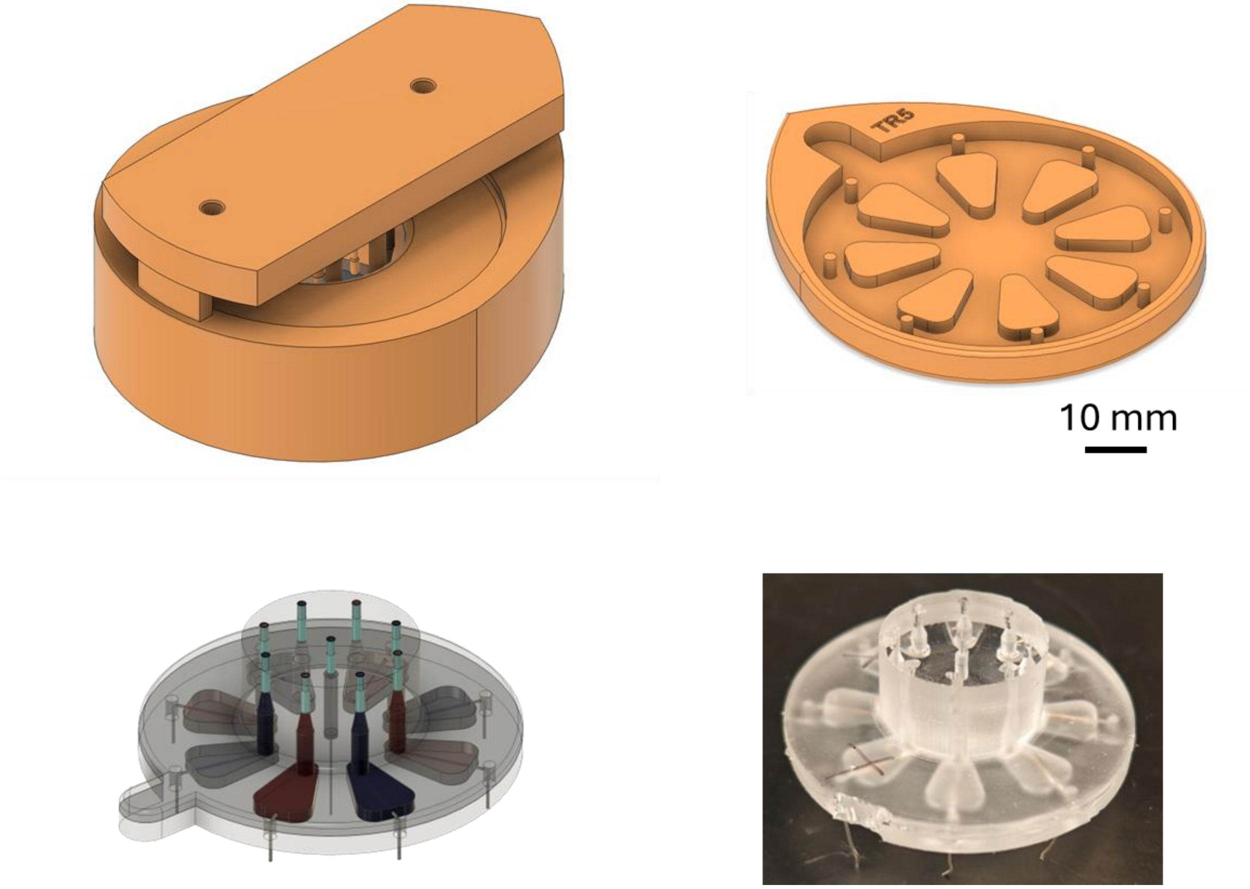


**Fig. S3.**
Molding of PDMS body for wearable actuators.

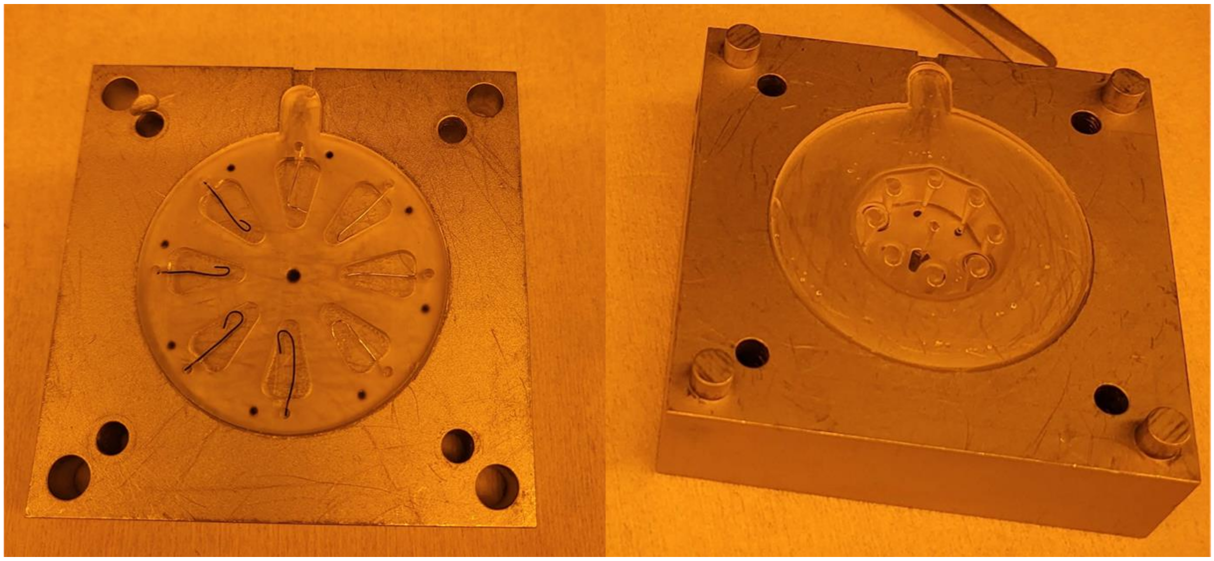

**Fig. S4**
Bonding of PDMS using oxygen plasma and aluminum clamps.

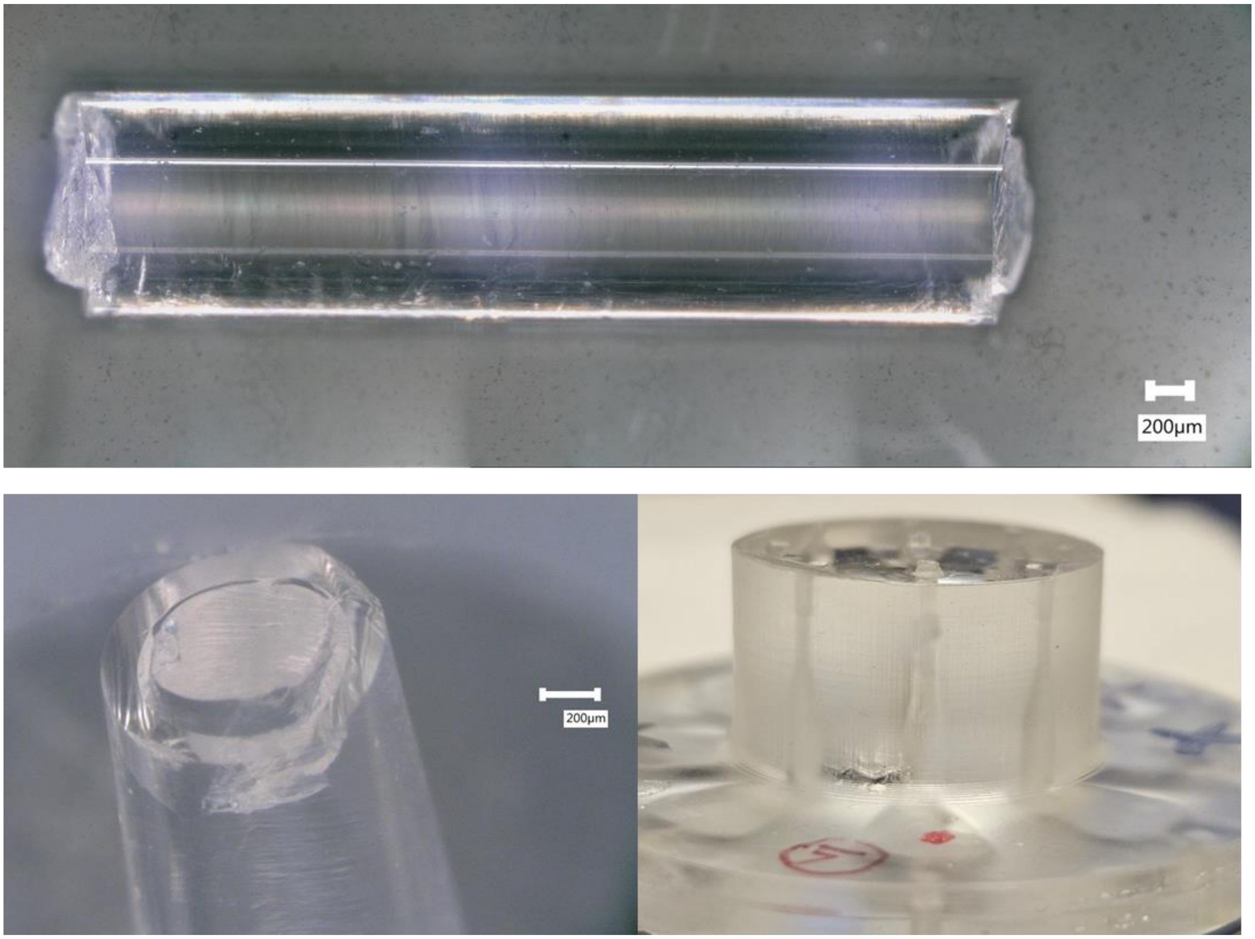


**Fig. S5.**
Microscopy image of the capillary with charge selective hydrogel.

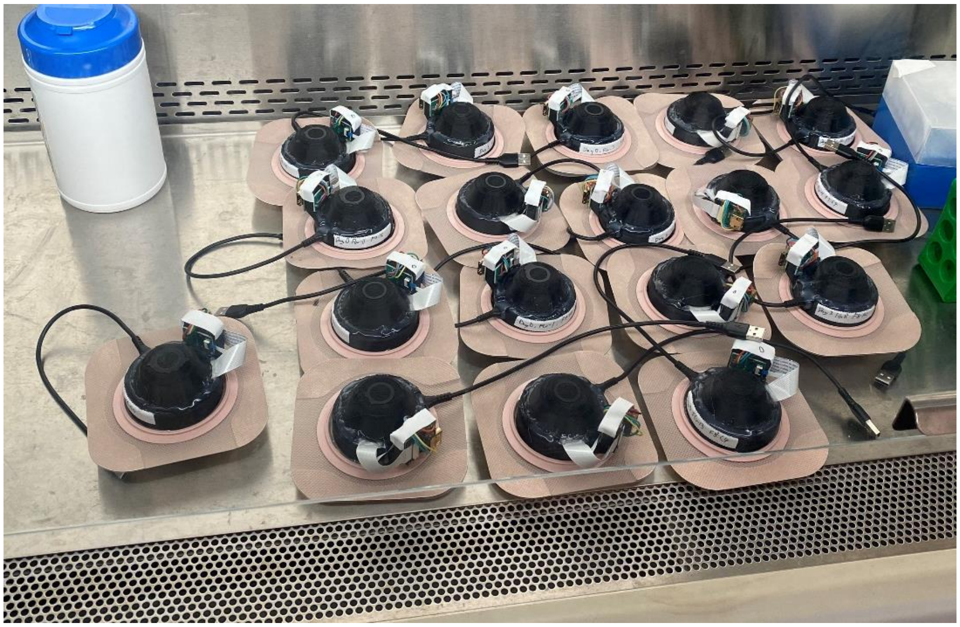

**Fig. S6**

A batch of fabricated Devices for ex vivo and in vivo tests in a sterile environment in a biosafety hood.

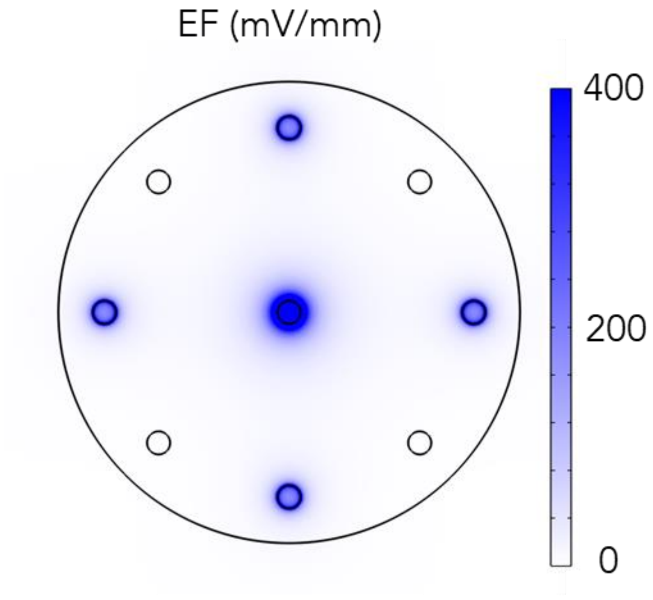


**Fig. S7.**
EF strength distribution using 400 mV/mm range, showing stronger EF near electrodes

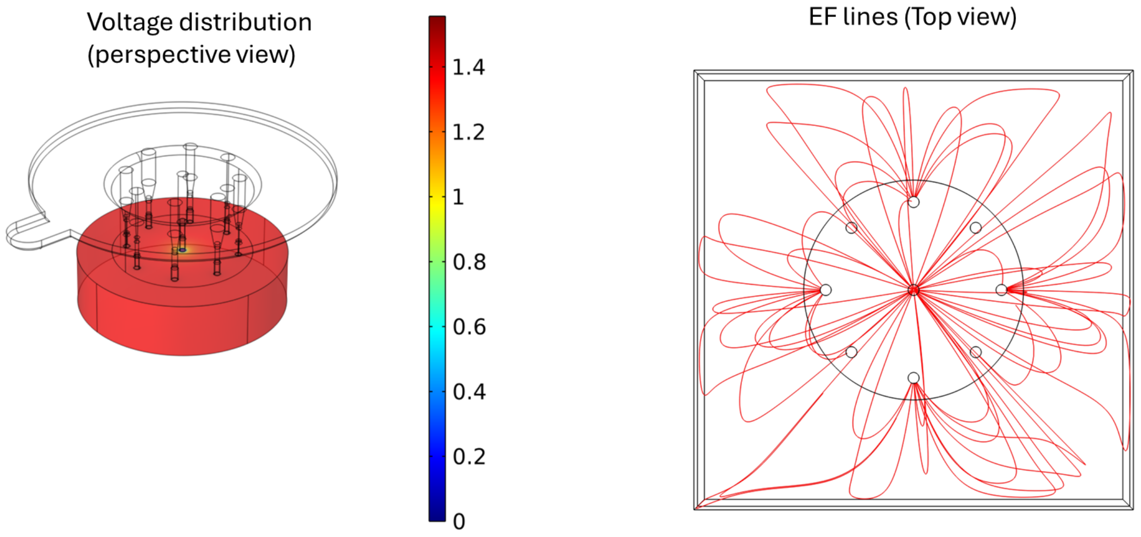


**Fig. S8.**
Comsol simulation of voltage distribution and EF lines. Simulation result using finite element simulation gives estimated resistance from edge channel to center channel to be 31 kohm.

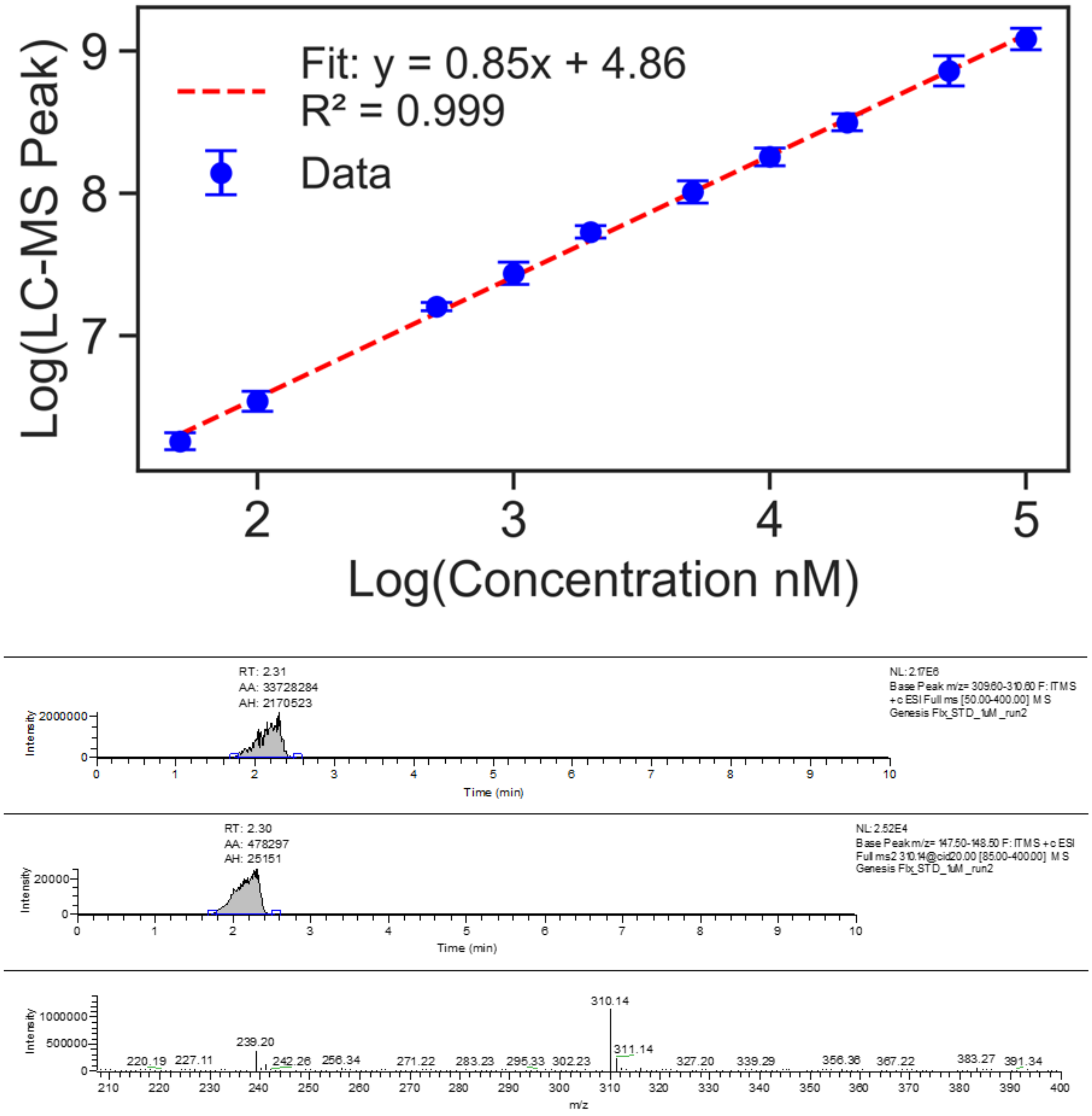


**Fig. S9**

HPLC-Mass Spectrometry calibration curve for measurement of Fluoxetine concentration. Peak at 310.14 corespondes to fluoxetine ion.

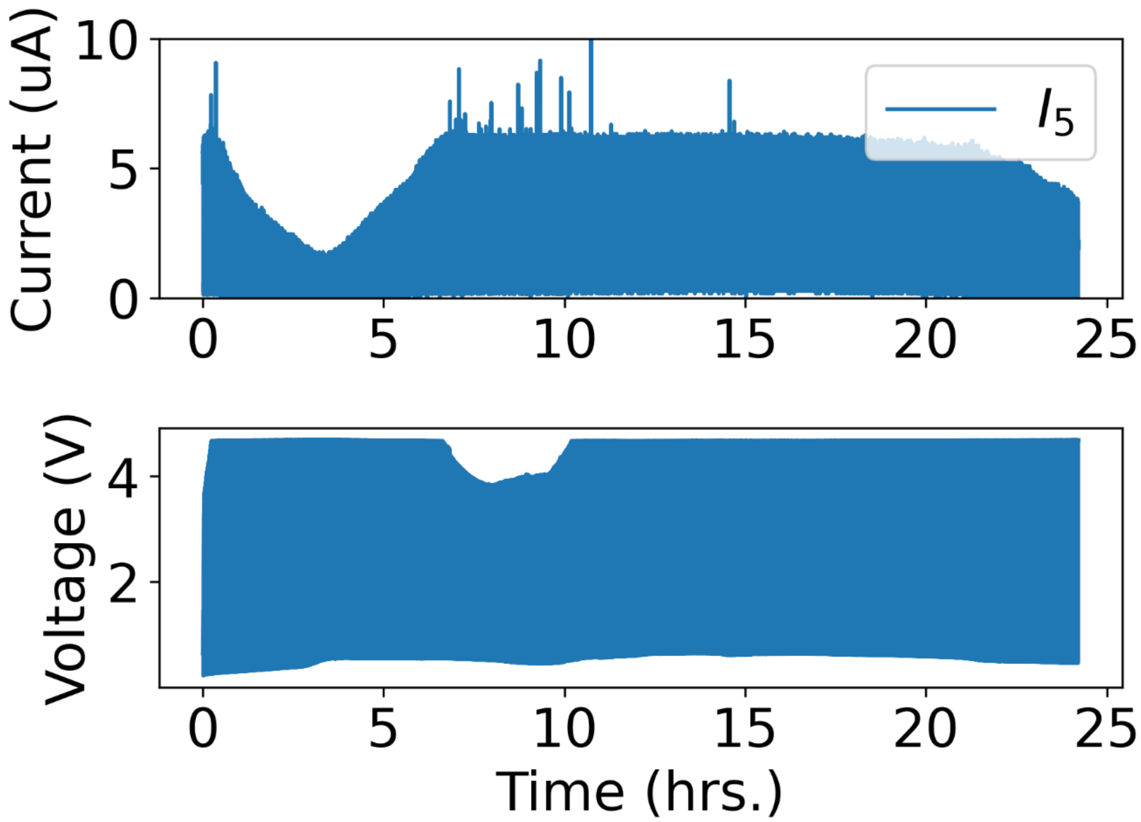


**Fig. S10.**
Current and Voltage of fluoxetine delivery through one channel for efficiency test.

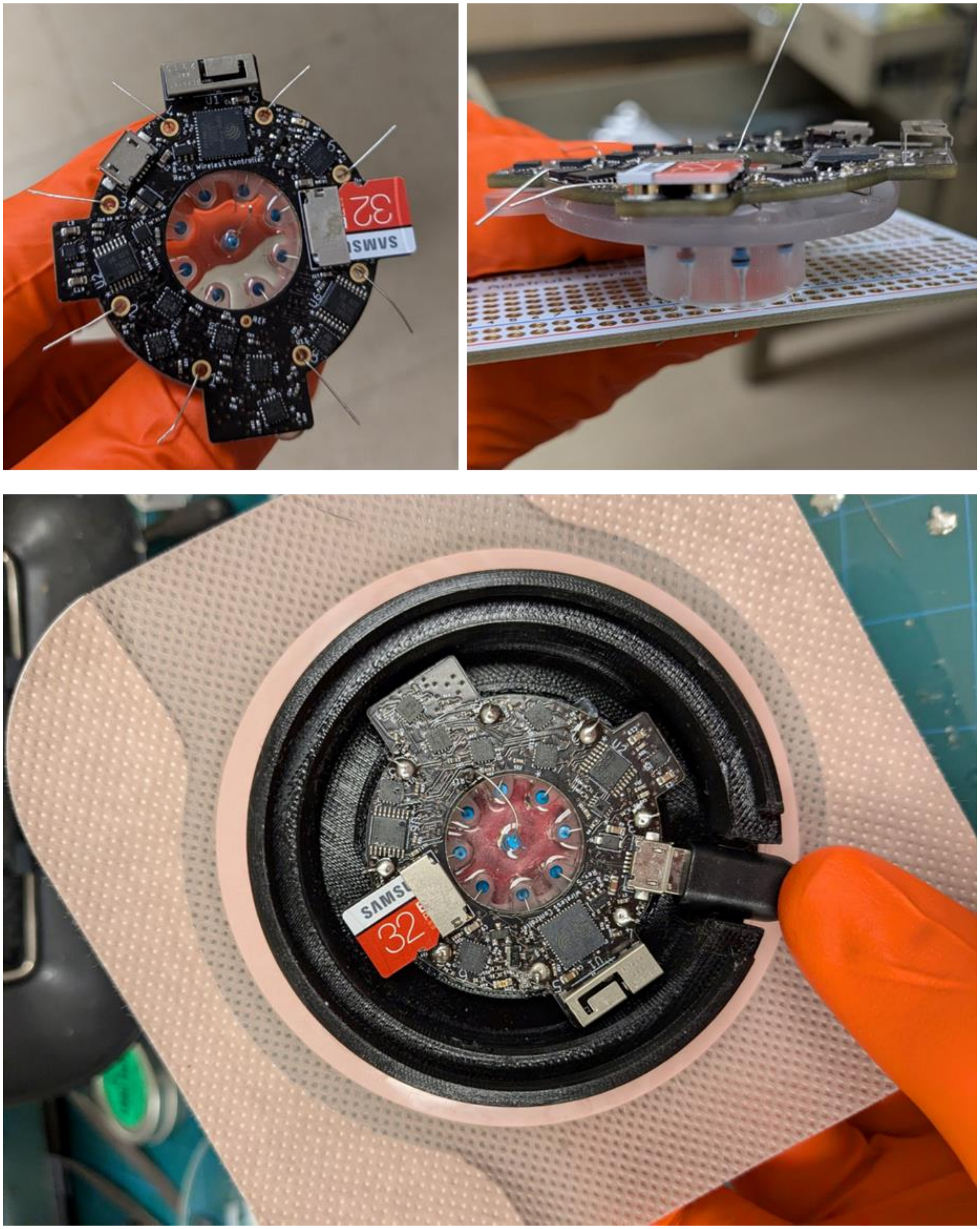


**Fig. S11**

Testing wearable device with resistors to validate the device performance.

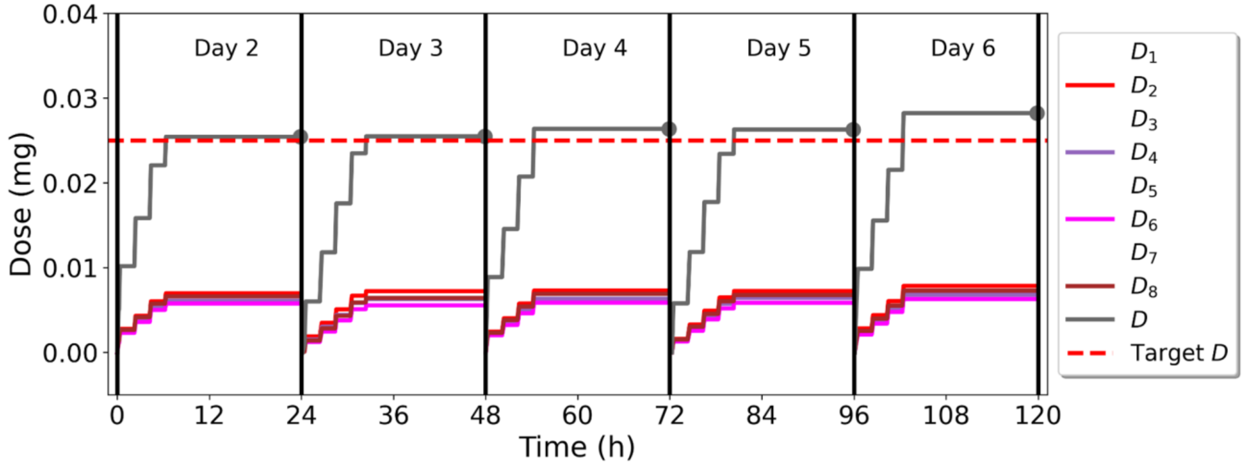


**Fig. S12.**

Test results using resistors to validate the cut-off at a daily maximum dose of 0.025 mg/wound.

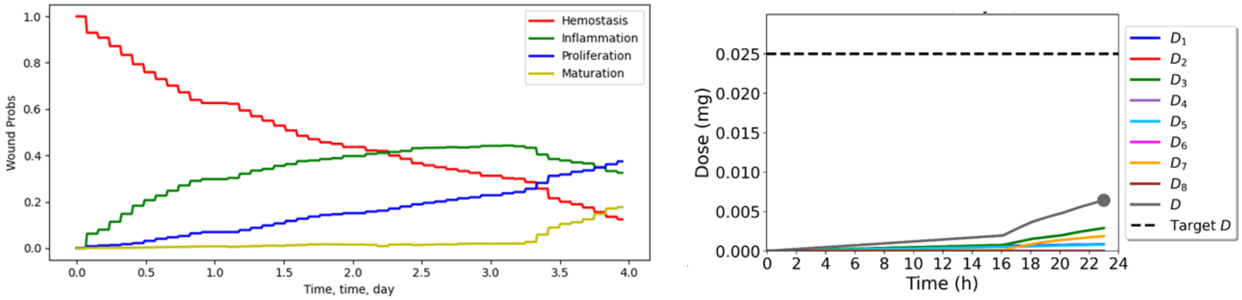


**Fig. S13.**

Algorithm-based switching between EF and Flx treatment in dummy test in vitro using resistors and pre-recorded wound images.

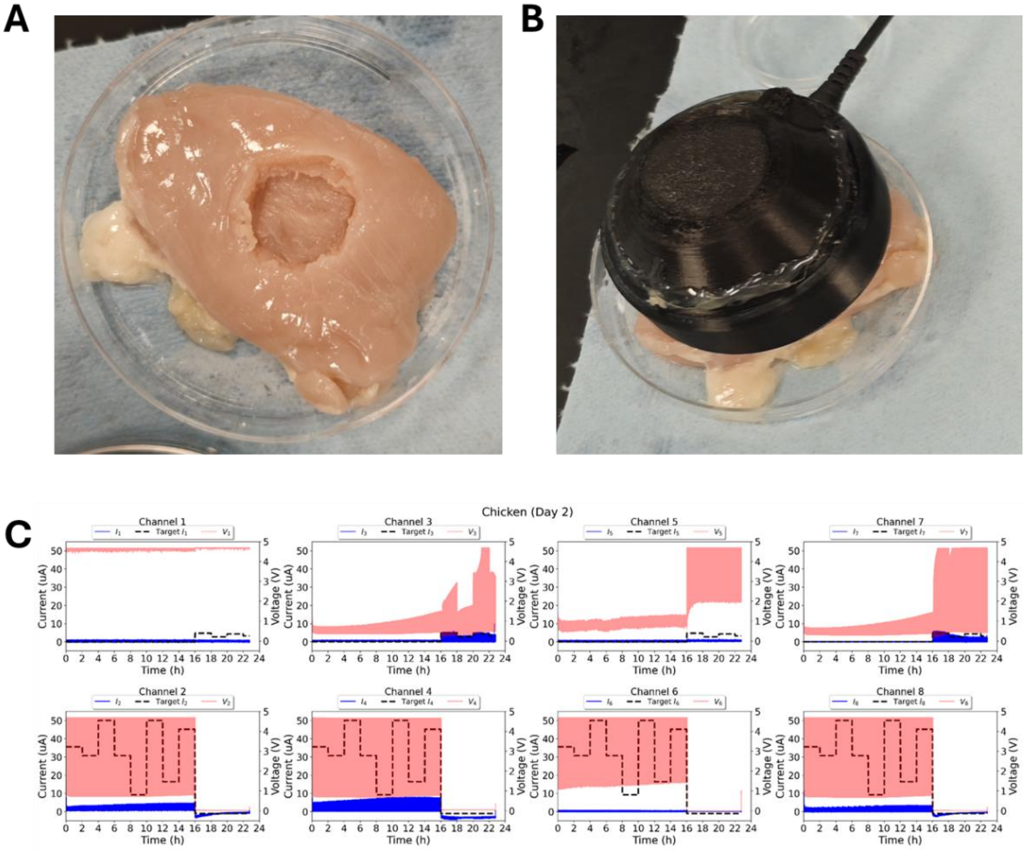


**Fig. S14.**

Test of device performance ex vivo. (A)20 mm dummy wound created chicken breast. (B) Device mounted on dummy wound. (C) Device performance with 8 channels. Even channels were turned on first then odd channels were turned on after the switch event at 16 hr.

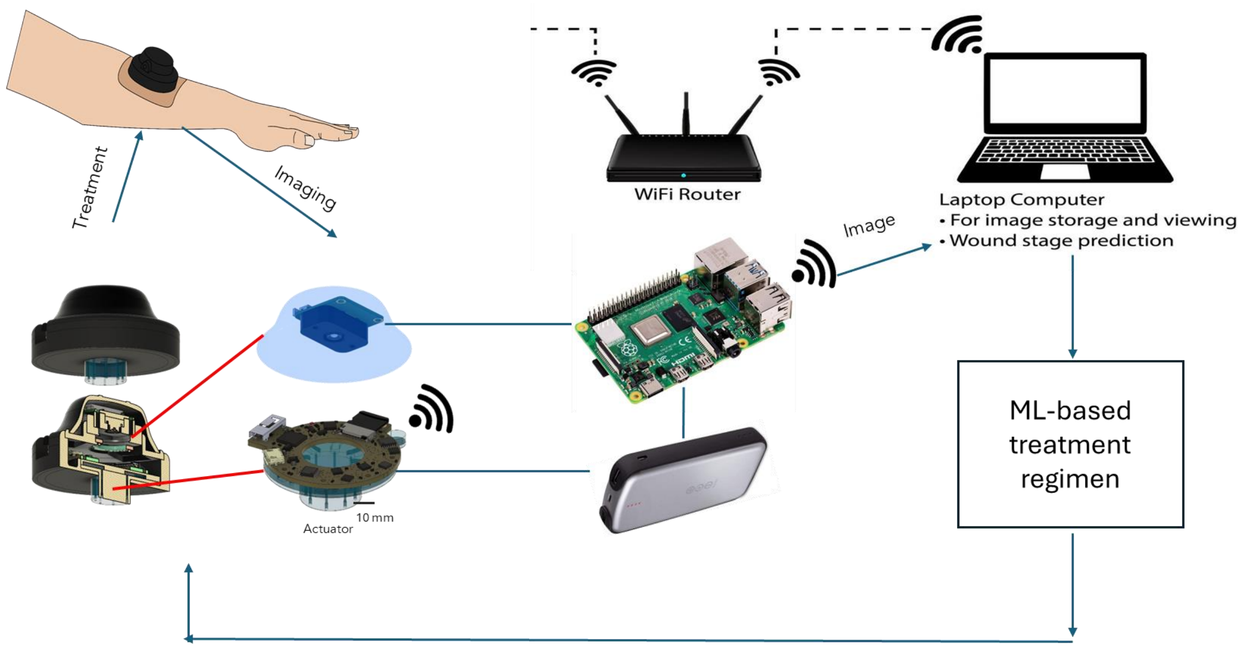


**Fig. S15.**
Set up of connection of the system

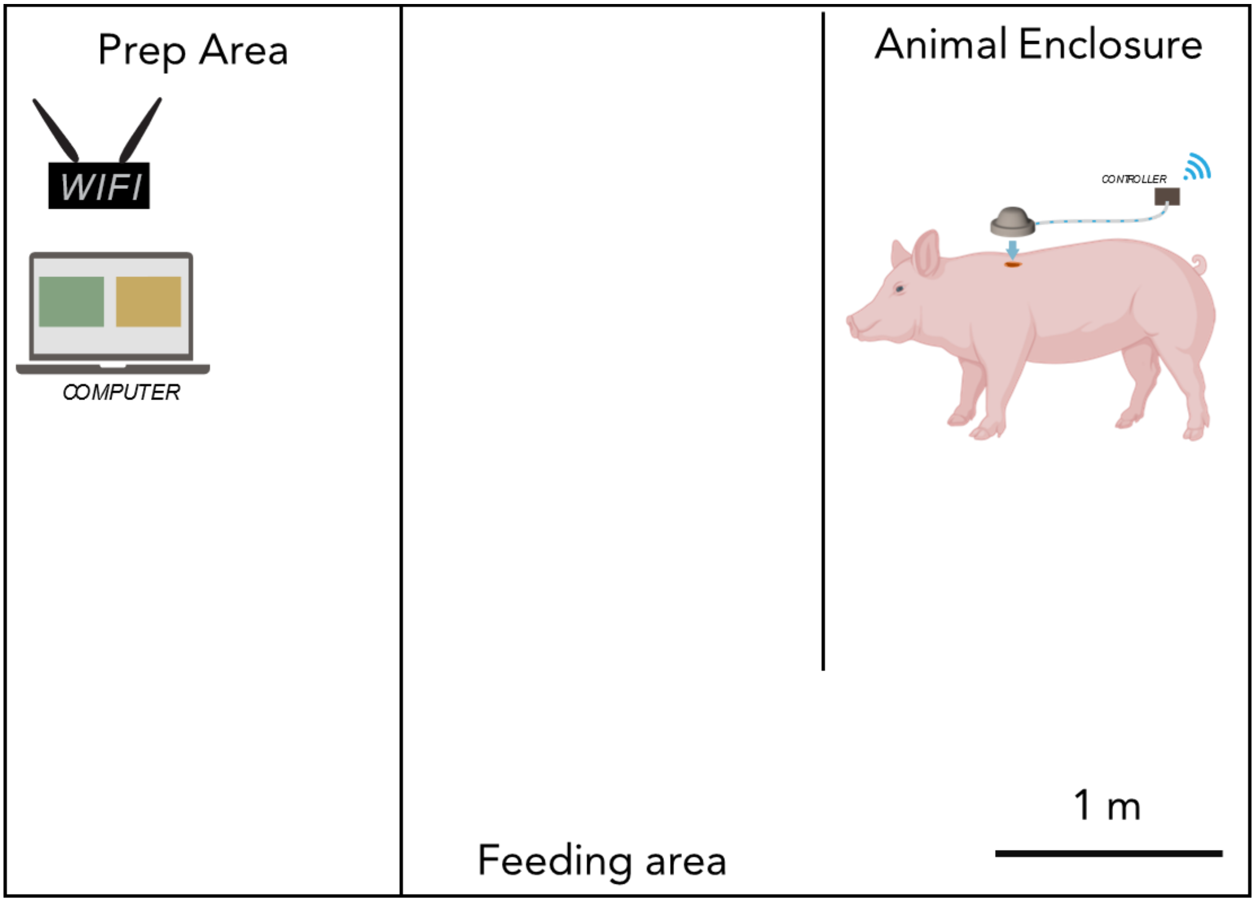


**Fig. S16.**
Set up of router and computer in the animal vivarium.

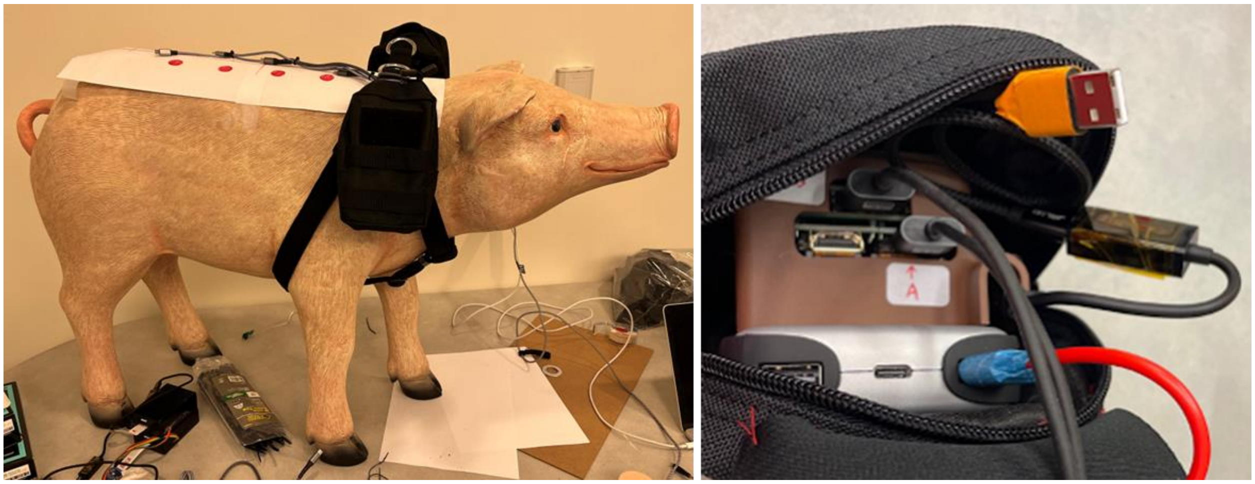

**Fig. S17.**
Setup of pouch on harness that contains battery and Raspberry Pi.

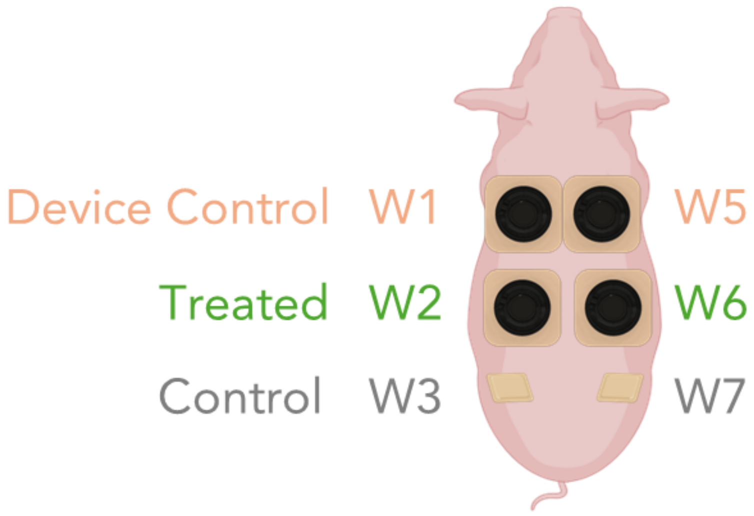


**Fig. S18. Wound map of the porcine model.**

W1 and W5 are device control without actuation. W2 and W6 are wounds treated with TheraHeal system, W3 and W7 are Standard-of-care controls. W4 was skipped(no wounding) to keep numbering consistent across experiments.

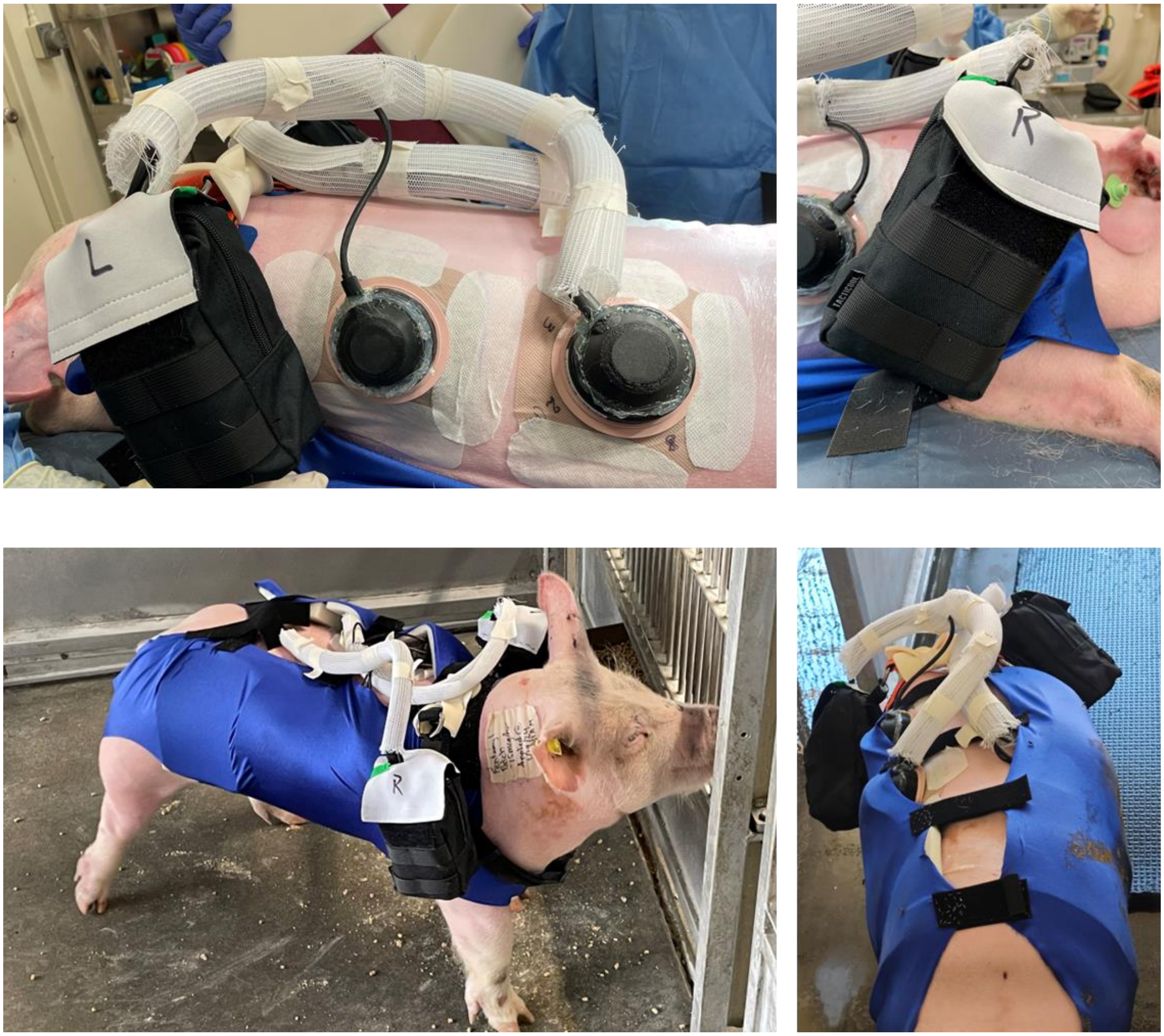


**Fig. S19.**

Porcine that mounted with the bioelectronic device, harness system with pouch.

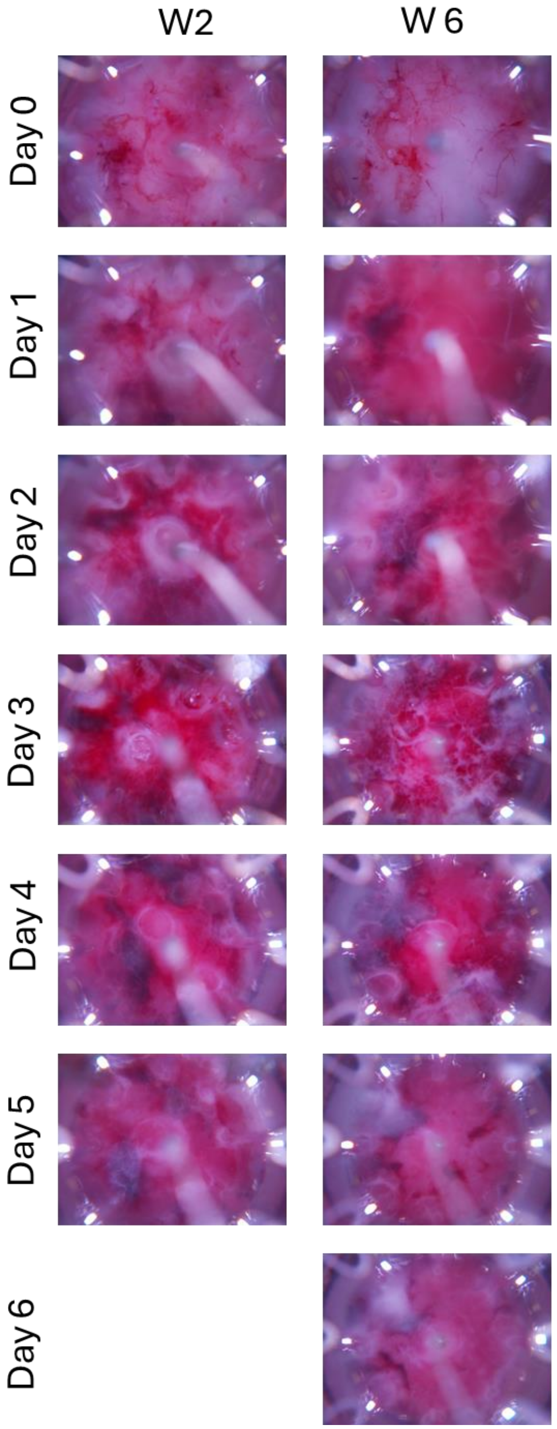


**Fig. S1**

picture taken by the wearable camera module for wound stage monitoring and evaluation.

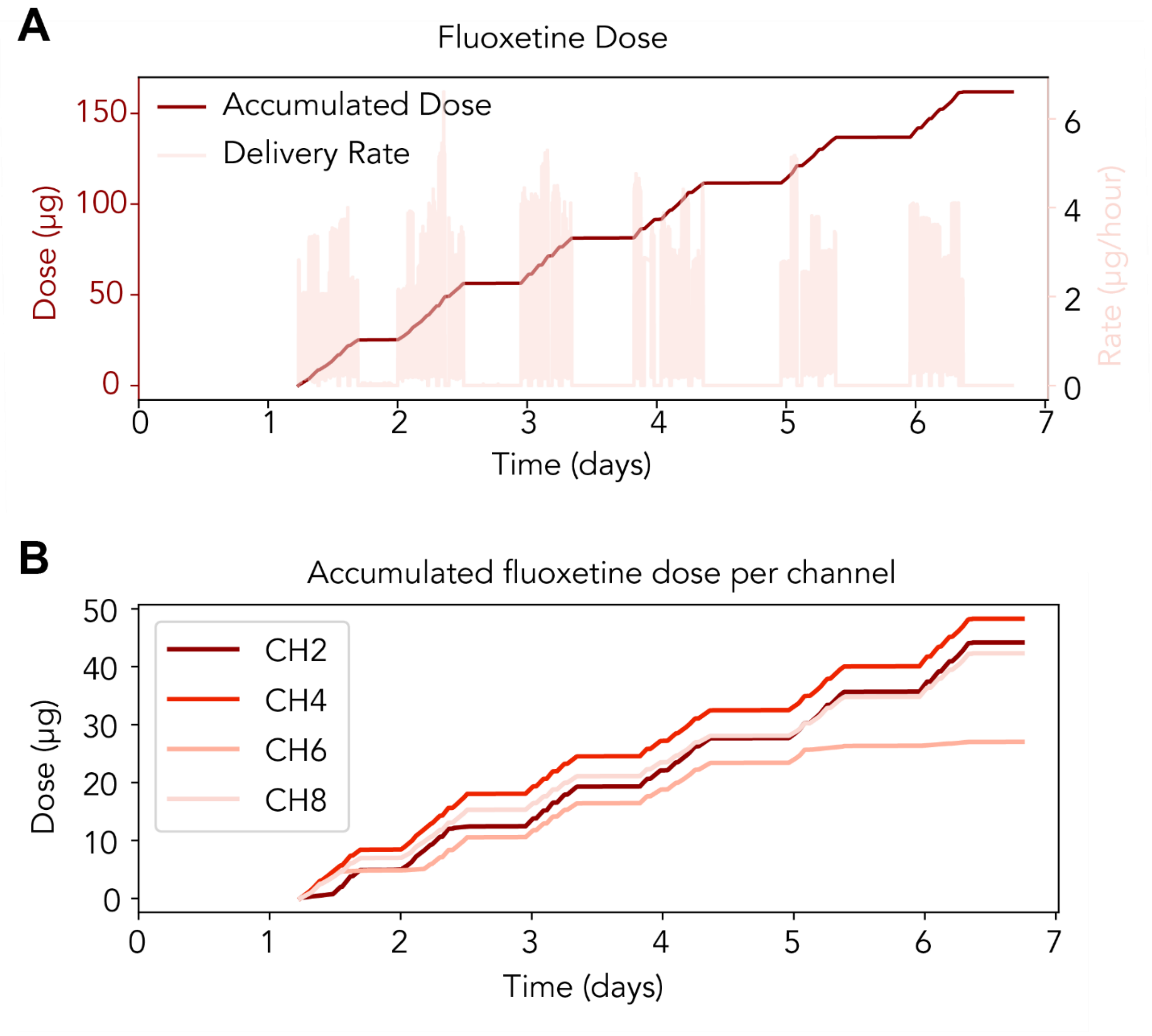


**Fig. S21. Accumulated fluoxetine dose.**

(A) Dose(left axis) and deliver rate (right axis) from the whole device with multiple channels.
(B) Dose from each individual channe.

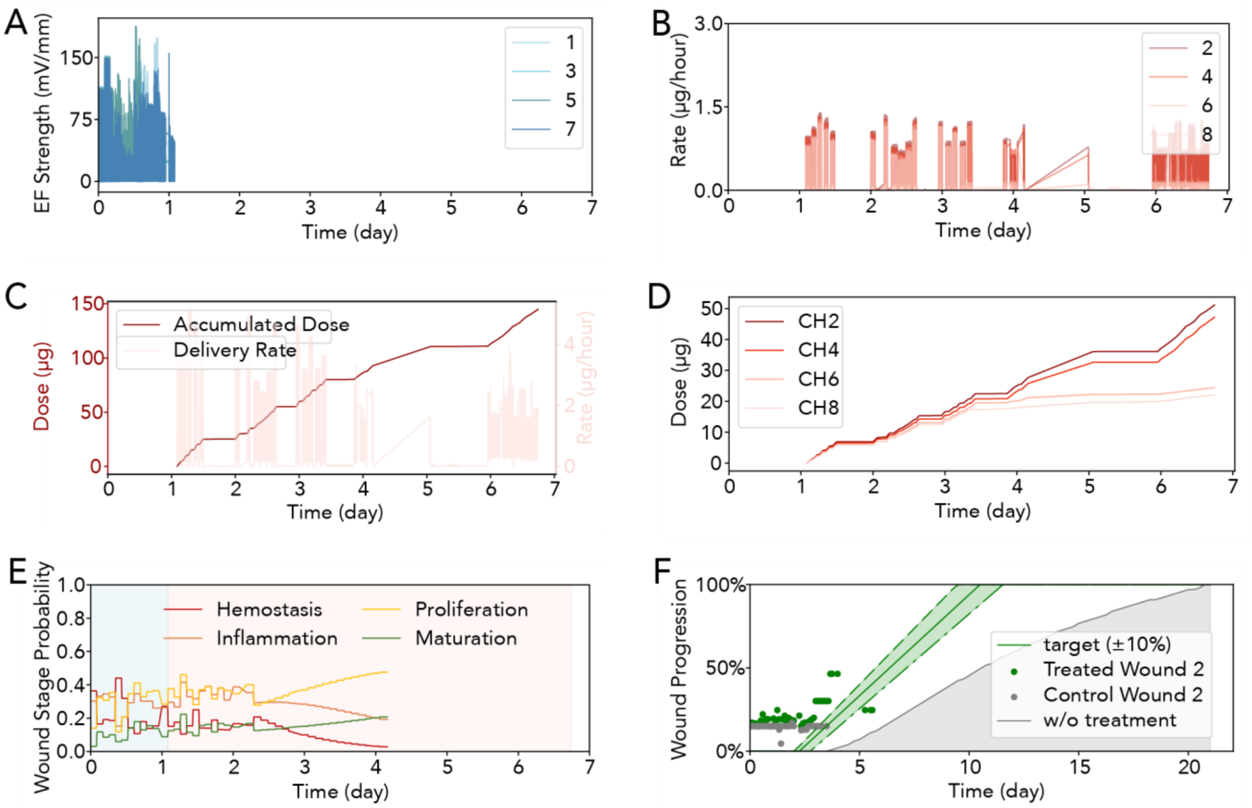


**Fig. S22. Treatment and analysis on wound W2**

(A) Electric field strength from individual channels of a bioelectronic device. (B) Fluoxetine delivery rate from individual channels of a bioelectronic device. C) Dose(left axis) and deliver rate (right axis) from the whole device with multiple channels. (D) Dose from each individual channel. (E) wound stage probability of a treated wound, interpreted by Deep mapper (F) Wound healing progress from control and treated wound, interpreted by Deep mapper.

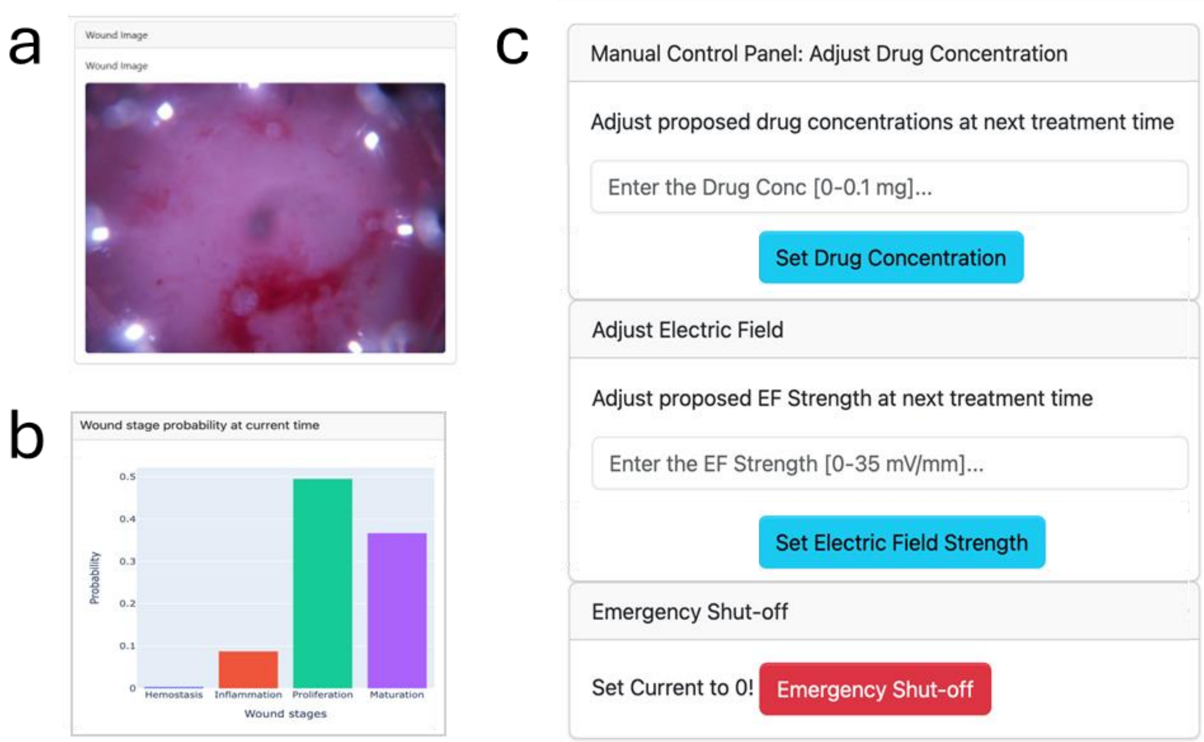


**Fig. S23**

Web based GUI to allow physicians to monitor the wound healing process and fine-tune treatments as needed in a clinical setting.

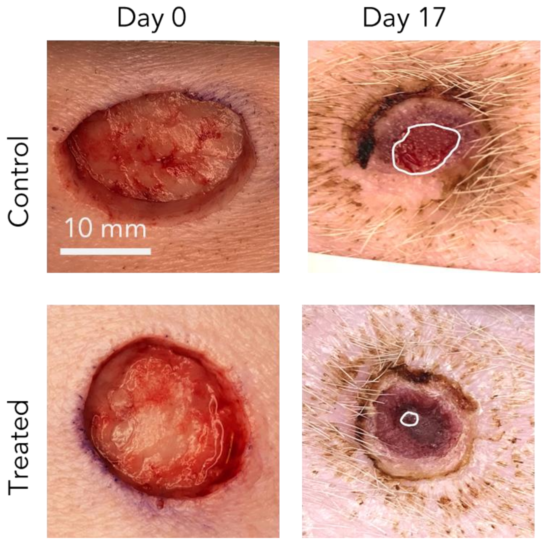


| Wound Area | | |
|---|---|---|
| | Day 0 | Day 17 |
| Wound 3 Control | 320.7 $mm^2$ | 32.7 $mm^2$ |
| Wound 6 Device | 354.6 $mm^2$ | 2.4 $mm^2$ |

| % of Day 0 Wound | | |
|---|---|---|
| | Day 0 | Day 17 |
| Wound 3 Control | 100% | 10.2% |
| Wound 6 Device | 100% | 0.68% |

**Fig. S24.**

Wound size evaluation from camera image. All wounds were created with 20 mm diameter and 6 mm thick at day 0; treated wound show smaller size at day 17.

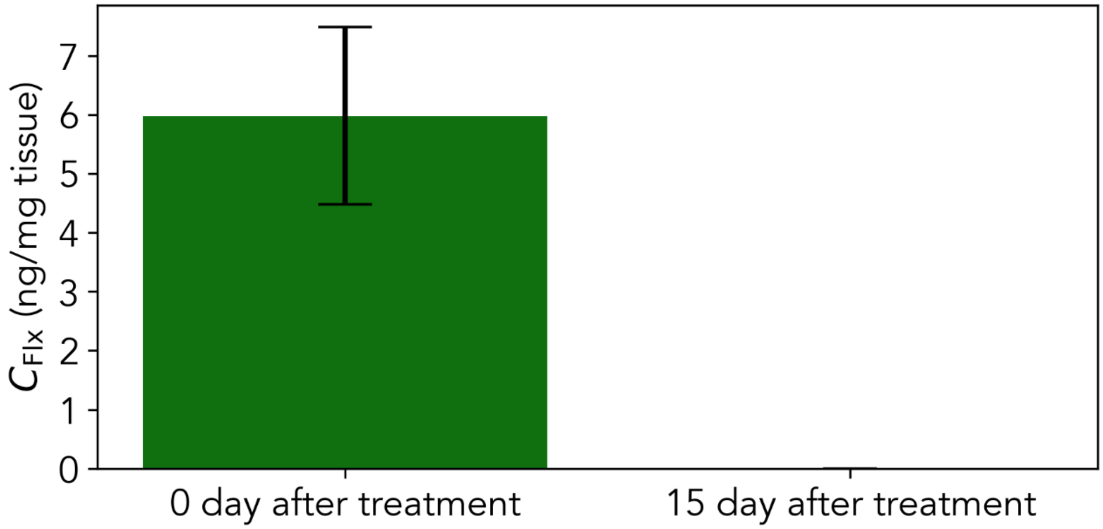


**Fig. S25.**

fluoxetine concentration in wound tissue following application using the experimental device. Topical fluoxetine was delivered to the pig wounds daily over the course of six to seven days. Day 7 of healing, n=8 wounds; day 22, n=2 wounds. Error bars show standard error.

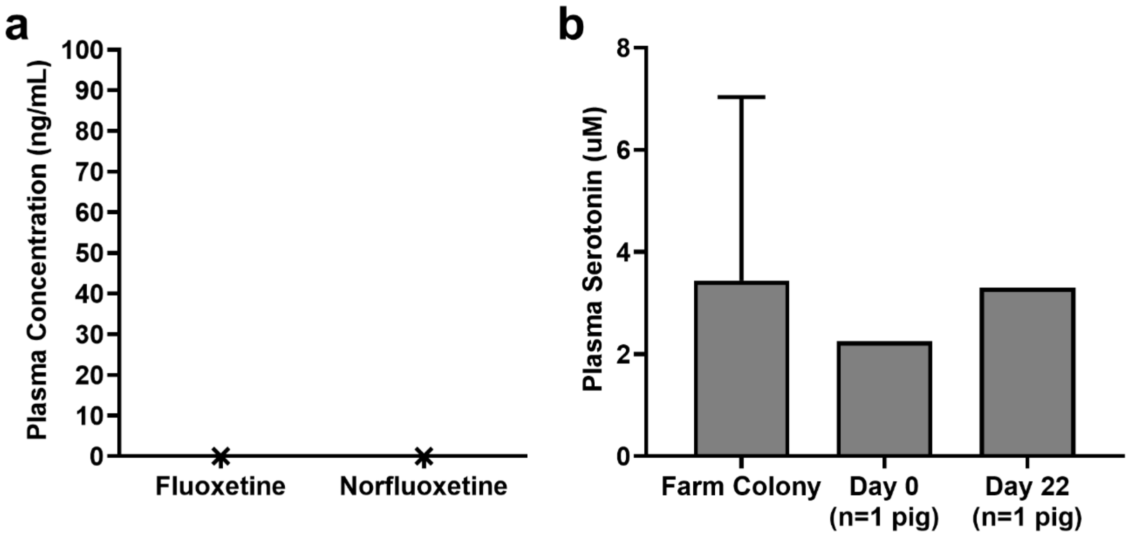


**Fig. S26.**

(a) Plasma concentrations of fluoxetine and norfluoxetine in one pig following treatment using the experimental device. (b) Normal plasma serotonin concentrations for the UC Davis farm colony (n=13), and plasma serotonin concentrations from our experimental animal at baseline and following treatment using the experimental device (n=1). Plots show mean ± SD.

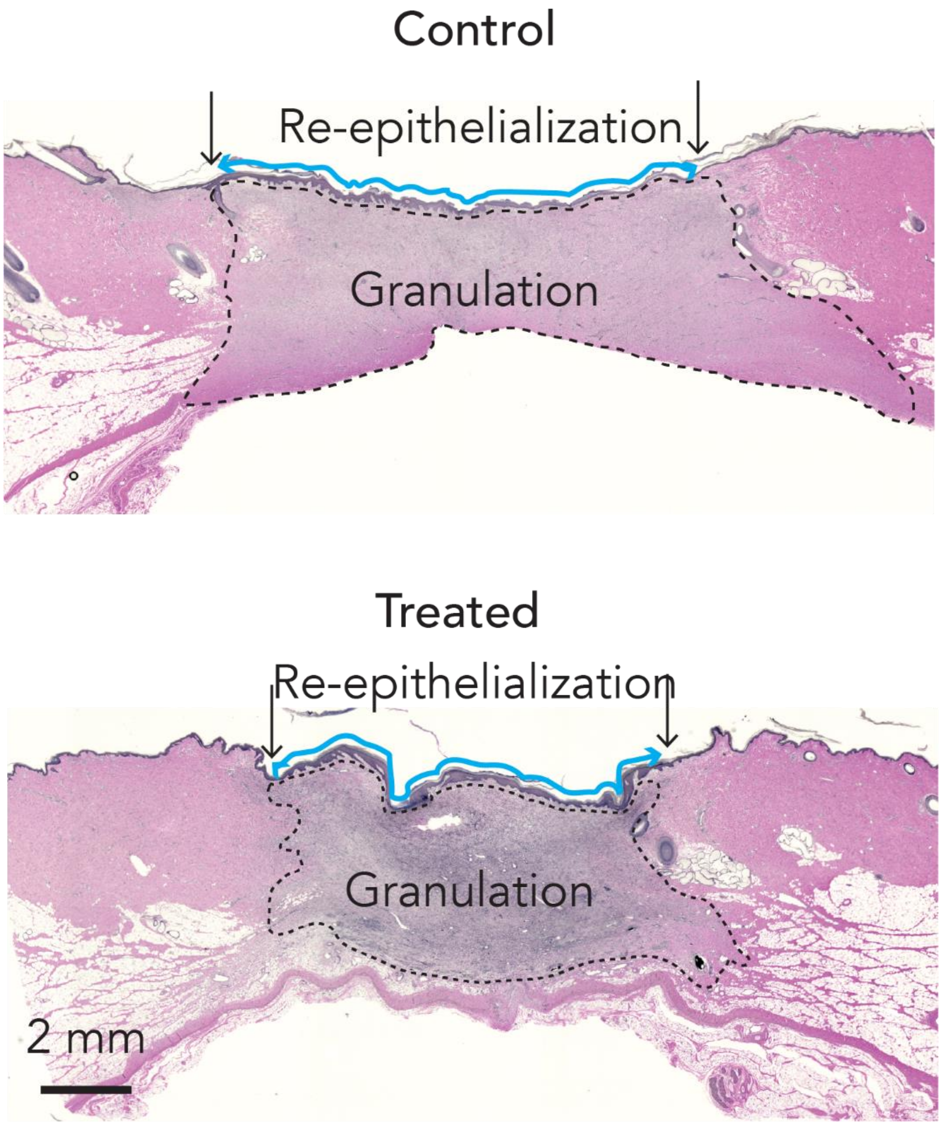


**Fig. S27.**

Representative images from the day 22 wounds treated with standard care or the combo device. The wound edge in each image is indicated by black arrows, and the re-epithelialization is shown by the blue lines extended on top of the regenerated granulation tissue (circled by the dotted line).

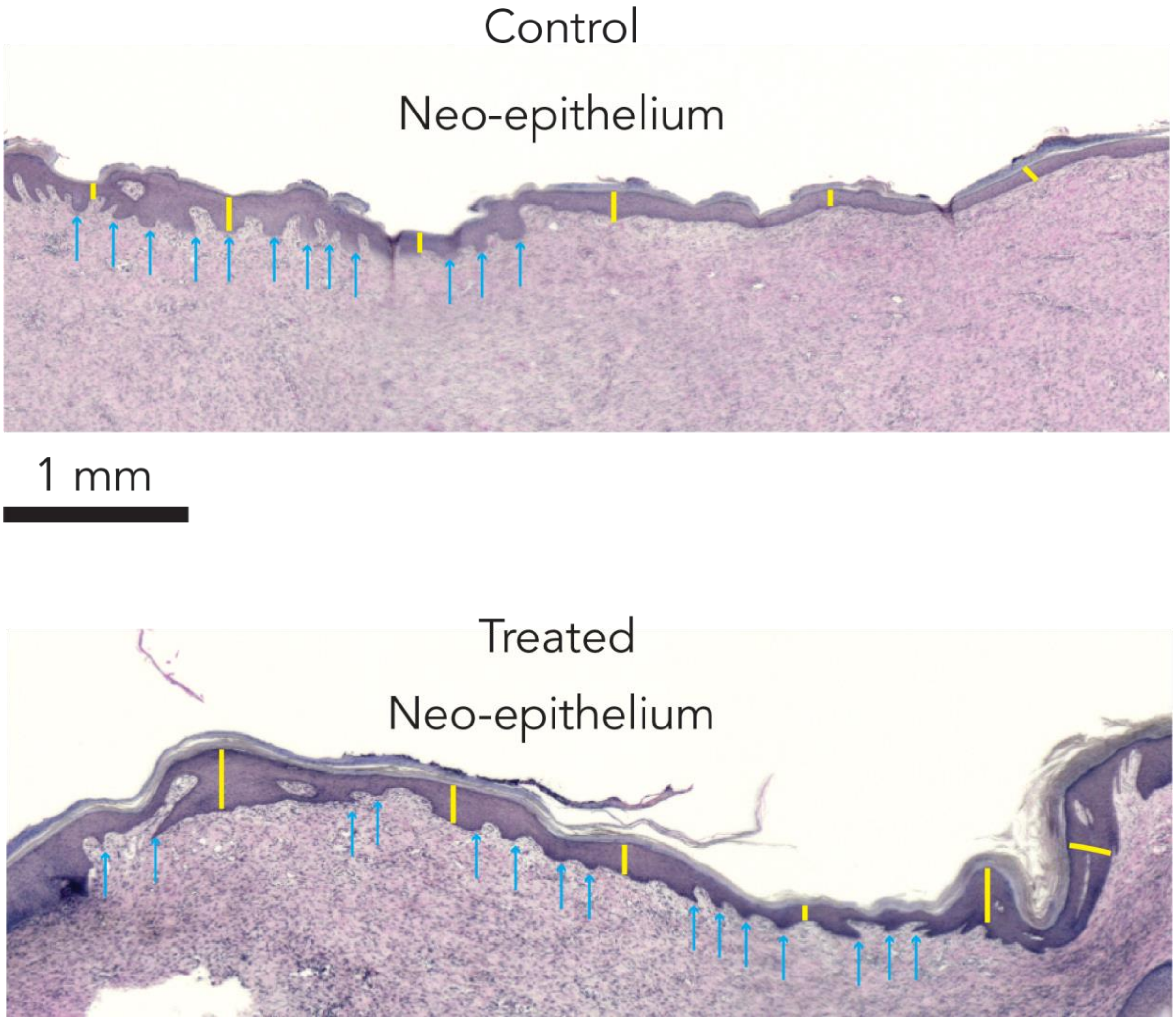


**Fig. S28.**

Measurements of epithelial thickness. Representative images from the neo-epithelium covering the re-epitheliaized wound on day 22. The epithelial thickness was measured from 10-12 spots across the neo-epithelium.

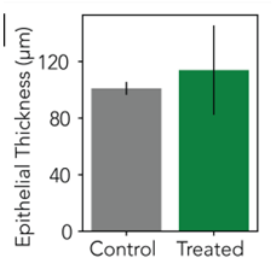


**Fig. S29.**

Measurements of epithelial thickness with W2 and W6 combined as treated group compared to standard of care.

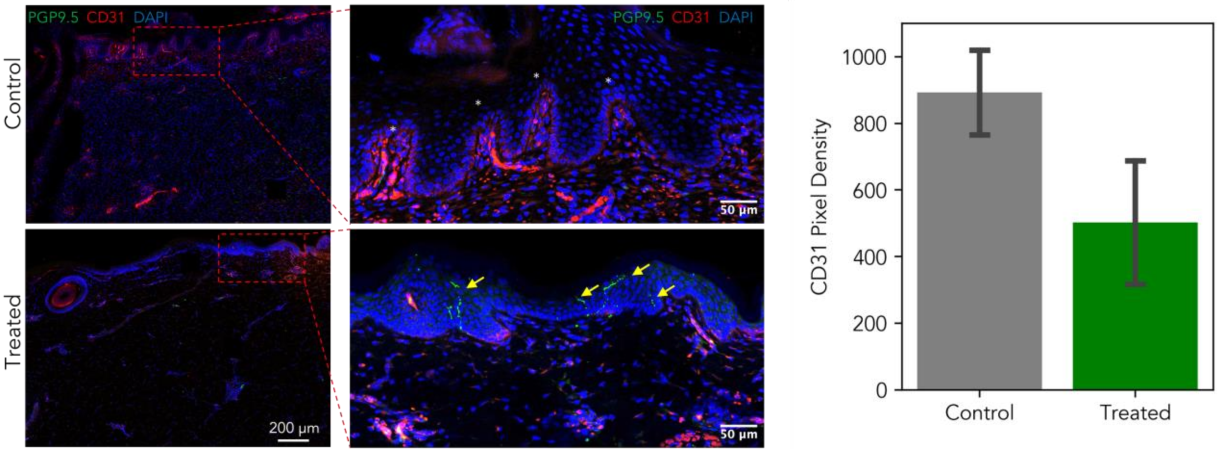


**Fig. S30.**

IHC staining of wound tissue after TheraHeal treatment. PGP9.5(green) staining Arrows show Nerve fibers in the fully healed epidermis. CD31 staining shows angiogenesis, Asterisk shows Granulation tissue with enriched vessel sprouts. Less angiogenesis indicated the later wound healing stage. Bar chart shows a 26% decrease in angiogenesis marked by CD31 pixel density (n = 2, p = 0.15)

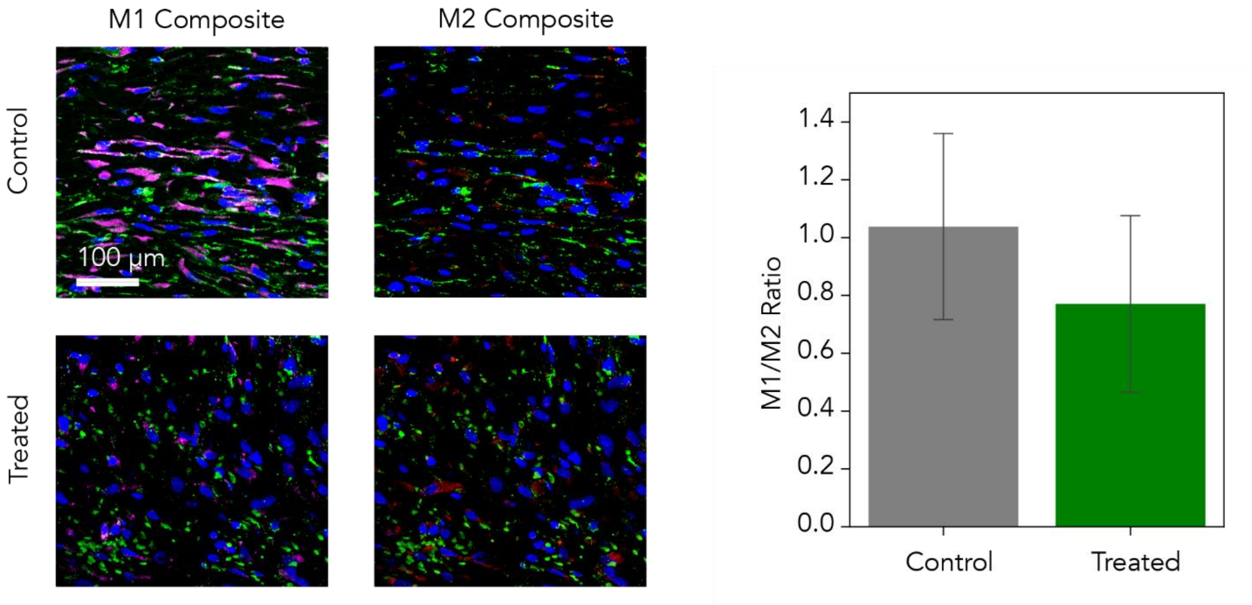


**Fig. S31.**

M1/M2 macrophage staining on control and treated wound from tissue collected on day 22. immunohistochemistry (IHC) staining to visualize macrophages on control and treated wounds from tissue collected on day 22. Macrophages were labeled with antibodies against iNOS (M1, purple), arginase 1 (M2, red), and a pan-macrophage marker (BA4D, green). Bar chart shows a 26% decrease in M1/M2 ratio($n = 6$, $p = 0.17$)

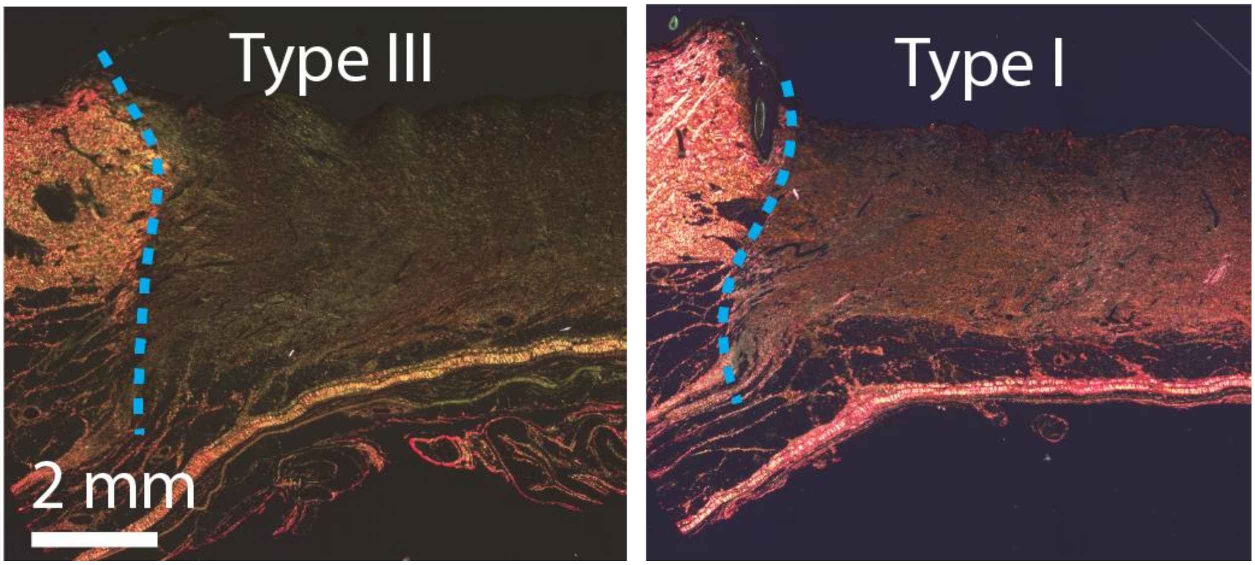


**Fig. S32.**

Representative polarized images of Picrosirius Red staining of the wound tissue, that selectively identifies  Type I and III collagen. Blue dotted lines indicate the wound edge.

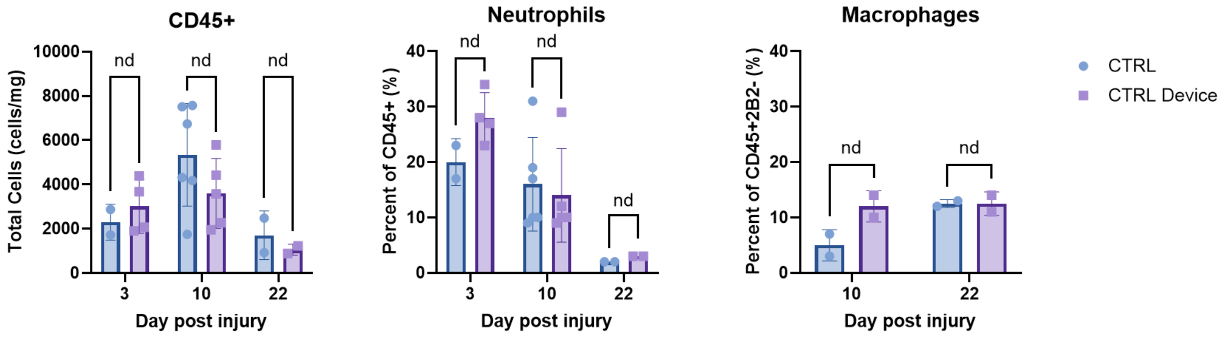


**Fig. S33.** Flowcytometry data of tissue sample. Control devices show no difference with the standard of care in immune cells (CD45+), macrophages (CD45+2B2-BA4D5+), or neutrophils (CD45+2B2-), indicating that the device interface itself won't trigger inflammation or immune response.

**Table S1. LC-MS measurement of delivery efficiency of Flx ion pump.**

| Sample | LC-MS peak Area | C(nM) | nmole | diffusion nmole | Coulomb | Net eff | on off ratio |
|---|---|---|---|---|---|---|---|
| Dev1 | 1.45E+06 | 2616 | 20.9 | 8.4 | 0.047 | 2.55% | 2.5 |
| Dev2 | 1.68E+06 | 3029 | 24.2 | 11.2 | 0.036 | 3.54% | 2.2 |
| Dev3 | 5.23E+07 | 2283 | 18.3 | 9.5 | 0.031 | 2.72% | 1.9 |
| Dev4 | 2.21E+07 | 833 | 6.7 | 1.7 | 0.343 | 0.14% | 3.9 |
| Dev5 | 9.30E+07 | 4474 | 35.8 | 14.2 | 0.101 | 2.06% | 2.5 |
| mean | | | | | | 2.2±1.1 % | 2.6±0.7 |

**Table S2. qPCR gene relative expression results on treated and control wound**

| Target Gene | Healthy Skin | Treated | | Standard of Care | |
|---|---|---|---|---|---|
| | | W6 | W2 | W7 | W3 |
| IL1b | 1.00 | 0.54 | 0.54 | 1.15 | 0.98 |
| IL6 | 1.00 | 3.73 | 4.60 | 2.77 | 3.81 |
| IL10 | 1.00 | 5.51 | 3.23 | 2.78 | 2.92 |
| IGF1 | 1.00 | 1.10 | 1.31 | 0.79 | 0.74 |
| TGFb1 | 1.00 | 8.70 | 5.71 | 5.87 | 5.05 |
| TNF | 1.00 | 0.47 | 0.43 | 0.36 | 0.46 |
| VEGF | 1.00 | 1.25 | 1.30 | 0.97 | 1.14 |

**Data S1. (separate file)**

CAD design of wearable devices.

**Data S2. (separate file)**

Biocompatibility report from Da Vinci Biomed Research Product, inc.
